# Protecting Episodic Memory After Sleep Loss: Similar Benefits of Exercise and Naps via Distinct Neural Contributions

**DOI:** 10.1101/2025.08.19.25333837

**Authors:** Madhura S Lotlikar, Beatrice Ayotte, Amy Choi, Freddie Seo, Edwin M Robertson, Fabien Dal Maso, Marc Roig

## Abstract

Sleep benefits episodic memory, which is critical for everyday cognition, future planning and decision-making. Sleep loss, a widespread issue across all ages and a major public health concern, impairs the brain’s capacity to encode episodic memories. This, in turn, disrupts cognitive functions that rely on episodic memory, such as decision-making based on past experiences, encoding new events, or recalling critical safety protocols posing risks, particularly in safety-sensitive occupations. Cognitive impairment due to sleep loss at such workplaces raises safety concerns and causes accidents. Merely implementing sleep hygiene techniques may not be effective or practical in such settings. Finding cost-effective strategies to preserve episodic memory after sleep loss is critical. We compared the effect of exercise and naps to reduce the impact of sleep loss and investigated mechanisms underlying their potential benefits.

Fifty-four healthy young (18-35 years) individuals were subjected to 30 hours of continuously monitored wakefulness after which they were randomized into a 90-minute nap (NAP) (n= 18), 20-minute exercise (EXE) (n= 18) or do nothing (Control: CON) (n= 17, 1 excluded) groups. Following this, all participants were shown images (encoding), and three days later their memory was tested in a Yes-No recognition paradigm by presenting a mix of previously shown images and new ones. Electroencephalography (n= 43) from the encoding session was analyzed for pre-stimulus sleep pressure and fatigue markers: delta/theta spectral power; and episodic memory encoding markers: event-related beta desynchronization (beta-ERD), event-related delta/theta synchronization (SW ERS) and P300 component of event-related potential.

Both EXE and NAP groups had higher memory for the encoded images than the CON group (Cohen’s d 1 and 0.91, respectively; with average improvements of 22% over the CON group), and both intervention groups had similar memory scores. Contrary to the literature in normal wakefulness, beta-ERD and P300 amplitude did not differ significantly between EXE and CON groups, and only in the EXE group these two markers were associated with memory. In the CON group, in contrast, P300 was associated with fatigue. While all the groups showed delta and SW ERS, only in the NAP group were these markers associated with memory. Regression analyses revealed that the best neural predictors of memory performance in the EXE group were P300 and beta-ERD on remembered trials (Rsq. adj. 0.64). In contrast, in the NAP group, memory performance was best predicted by sleep pressure markers and SW ERS on remembered trials (Rsq. adj. 0.77). None of these predictors explained memory performance in the CON group.

In summary, we demonstrate that exercise and napping benefit episodic memory performance after sleep loss with equal magnitude, but through different neural contributions within each group. Under a sleep-deprived state, exercise facilitates efficient neural processing while napping makes the brain state conducive to new learning, which contributes to memory encoding. Our mechanistic findings strengthen the principle of neural degeneracy. These results have important societal and policy implications for preserving performance under sleep-deprived conditions.

## MAIN TEXT

A staggering 20 – 40% of adults worldwide sleep insufficiently and ∼8.2 million people in Canada work night shifts, many in high-stake safety-sensitive occupations, and face sleep disturbances and insufficiency^1–4^. Sleep insufficiency increases the risk of various neurological and psychiatric conditions, substance abuse, all-cause mortality, traffic-related and industrial accidents, medical errors, decreases workplace productivity and jeopardizes public safety^4^. Altogether, about $680 billion is lost per year across five OECD nations due to insufficient sleep, making it a growing public health concern^4^. This pervasive nature of sleep insufficiency and the limited time availability during shifts for essential workers makes it urgent to explore non-pharmacological, acute interventions, which can mitigate the negative impact of sleep loss on the brain and different aspects of cognition such as episodic memory^5^. Indeed, it’s known that sleep contributes to episodic memory by facilitating not only the consolidation of previously learned information but also the encoding of new information^6–9^. In contrast, a night of sleep loss impairs episodic memory encoding capacity^10–12^.

Episodic memory is a type of memory that enables conscious recollection of past events and experiences within their spatiotemporal context, orients us in the present, and supports future planning and decision-making^13–16^. This memory plays a critical role in everyday cognition, especially in the high-stake safety-sensitive occupations - healthcare, law enforcement, transportation, and emergency services. These occupations demand optimal functioning of rapid and higher-order cognition relying on episodic memory, such as decision-making based on past experiences, encoding new events (e.g. surgical complications), and recalling correct safety procedures. Importantly, various brain regions such as the medial temporal lobe, frontal cortex, and sensory areas involved in episodic memory, also play crucial roles in other cognitive functions, including executive functions, attention, and information processing which are crucial^17,18^. Cognitive impairment impacting performance at such workplaces raises serious safety concerns and increases risks of medical errors, workplace injuries and accidents^1,4,19,20^. Thus, finding strategies to protect episodic memory and the neural systems supporting it after sleep loss is critical.

Addressing this issue requires multi-level strategies, as merely providing sleep hygiene techniques may not be effective nor practical^1,21–23^. In this study, we compared the effects of two widely accessible, low-cost, and side-effect–free interventions: an acute bout of cardiovascular exercise and a nap against a no-intervention control, on restoring episodic memory encoding capacity after a night of sleep loss. We also examined the neural mechanisms underlying the potential benefit using electroencephalography (EEG). Importantly, interventions were administered before memory encoding and not after because one night of sleep loss impairs memory more so when occurred prior to learning than after^24^.

While napping has shown to be effective in restoring alertness and alleviating fatigue and sleepiness and has also been recommended in some safety-sensitive occupations, nap opportunities could be restricted due to limited time availability, nap rooms, and implementation barriers^1,25–28^. Furthermore, naps may not be recommended in certain sleep disorders or when they are taken too close to bedtime^29,30^. This highlights the need to explore alternative interventions such as exercise, which may potentially offer similar benefits. To our knowledge, this is the first study to: a) examine the potential role of these two interventions to protect episodic memory from the effects of sleep loss, b) directly compare their benefits and c) investigate the underlying mechanisms under sleep deprived conditions. Beyond translational implications, our findings offer novel insight into how the sleep-deprived brain responds to two seemingly opposed neuroplasticity-modulating interventions benefiting memory such as exercise and napping^31,32^.

We chose these two interventions because they improve cognition and facilitate brain processes involved in episodic memory during normal wakefulness. Twenty minutes of moderate to vigorous intensity exercise performed before episodic memory encoding can improve long-term memory compared to not exercising^33,34^. This benefit may be mediated, at least in part, by increasing neurotransmitter release that facilitates long-term potentiation (LTP) - a synaptic mechanism of memory formation - in the hippocampus; or by priming neural networks to encode subsequent stimulus by inducing expression and post-translational modification of plasticity-related proteins; or by increasing neuronal excitability and enhancing attention^35,36^. Consequently, by promoting neuroplasticity, chronic exercise has been shown to counteract the deleterious effects of sleep deprivation on memory in animals^37^. Supporting this, human research has shown that people with high cardiorespiratory fitness, which can be increased by aerobic exercise training, tend to be less vulnerable to the deleterious effects of sleep deprivation on episodic memory^11^. Although plenty animal studies have shown the protective role of chronic exercise on memory after sleep loss, in humans few studies have explored the synergistic effects of exercise and sleep on memory, and no study has investigated the potential protective role of acute exercise on episodic memory after sleep loss^37,38^. Given the acute effects of exercise during normal wakefulness, we hypothesized that twenty minutes of aerobic exercise would prime memory encoding related neural mechanisms and benefit episodic memory under sleep-deprived conditions.

On the other hand, daytime naps ranging from 6 minutes to 120 minutes benefit alertness and mood in partially sleep-restricted conditions^30,39–42^. Importantly, after a night of sleep loss, 30 minutes and 60 minutes of naps failed to show the restorative benefits for vigilance and place-keeping tasks in a cohort of 280 participants, possibly because longer naps are required for higher-order cognitive functions, such as episodic memory^43^. This is supported by the evidence that slow wave sleep in naps is required to benefit declarative memories, and that nap duration is positively correlated with memory performance^12,44–46^. Based on this evidence, we hypothesized that 90 minutes of napping after sleep loss could renew the memory encoding capacity and benefit episodic memory performance. Together, we hypothesized that, compared to the control condition, both exercising and napping would protect memory encoding capacity from the effects of sleep loss and thus show higher episodic memory performance than the no-intervention control group.

Given the limited literature on neural mechanisms underlying episodic memory encoding under sleep-deprived conditions, we investigated the well-established state-dependent and stimulus-dependent markers of successful memory encoding under normal wakefulness conditions, with hypotheses informed by theories on memory function of sleep^9^. One such theory is the synaptic homeostasis hypothesis, which suggests that sleep renews the next day’s learning ability by globally downscaling synapses that have been potentiated throughout the wakefulness^9,47^. This is reflected as a wake-dependent rise and sleep-dependent decline in the molecular and electrophysiological markers of synaptic strength^47–49^. In humans, synaptic strength increase is reflected in a global, but predominantly frontal increase in waking slow wave oscillation/sleep pressure (delta and theta activity (0.5 - 8 Hz), which correlates with lapses during task performance^50–52^. Thus, compared to the nap group we expected higher levels of sleep pressure in the exercise and control group, which did not get an opportunity to sleep, and alleviate the sleep pressure. We also hypothesized that the sleep pressure will contribute to memory performance.

After sleep loss, in addition to an increase in sleep pressure, a larger local increase in delta and theta activity within task-associated regions can be observed before or after stimulus presentation which may result from the cumulative effects of the increase in sleep pressure, use-dependent local sleep pressure and task-associated fatigue^53,54^. This local increase has been associated with the lapses in executive function, attention, visuomotor task, and spatial navigation task performance^53,55–59^. Moreover, it is known that the pre-stimulus tonic brain state as indexed by an increase in slow wave oscillation in task-relevant regions contributes to the success of episodic memory^60,61,62^. Thus, we investigated whether these interventions could potentially benefit memory by preventing this local increase in slow wave oscillations during the pre-stimulus period thereby potentially mitigating the fatigue in task-associated areas and memory impairment. Beyond pre-stimulus state-related activity, we tested whether interventions could engage post-stimulus neural mechanisms of successful episodic memory encoding that are impaired in the sleep-deprived state. To this end, we first tested if the interventions modulated the event-related alpha/beta desynchronization (ERD) i.e. event-related oscillatory power decrease and theta/delta synchronisation (ERS) i.e. event-related oscillatory power increase during episodic memory encoding^63–66^.

It is well known that alpha/beta ERD correlates with neuronal firing rate reflecting active engagement of neurons and localized information processing while encoding an event^63,64,66,67^. The desynchrony allows the neuronal assembly to express a unique stimulus-specific code above noise, facilitating reliable information transfer, which is not efficient when the entire neuronal assembly is in synchrony^67^. Sleep loss impairs this capacity of beta desynchronization during hippocampal-dependent paired associates learning task^68^. In contrast, a study in older individuals showed that acute exercise increased ERD capacity which contributed to the cognitive benefit of exercise^69^. Thus, we hypothesised that the intervention groups will show greater beta ERD capacity than the control group, which will contribute to their memory performance.

Next, animal and human studies indicate that theta ERS supports item and context binding, enables temporal coordination of information flow between hippocampus and cortex and reflects neuronal dynamics optimal for LTP formation, thus supporting episodic memory^70–73^. Delta ERS has also been shown to be beneficial for various cognitive tasks such as episodic memory maintenance, mental calculation, working memory, semantic processing and inhibition^74–76^. Due to the limited evidence of ERS in the context of sleep loss, we hypothesized based on theory that the nap and exercise group would show higher delta/theta ERS (together referred to as slow wave oscillation ERS - SW ERS - hereafter) than the control group and will contribute to the memory performance. Importantly, we remained open to the possibility that interventions may engage distinct mechanisms to provide the memory benefit given their hypothesised differential effect on the brain state.

Finally, we compared the effects of the interventions on memory-related event-related potentials (ERPs). ERPs reflect time-locked voltage fluctuations of underlying neuronal assembly, which can capture the temporal evolution of cognitive processing of a specific event^77^. We conducted ERP analyses of P300 amplitude (positive ERP between 200-500ms after an event) in the task-relevant areas due to its contribution to memory and the influence of exercise on P300^78–82^. Briefly, the P300 component of the ERP signal indexes attentional resource allocation and inhibition of task-irrelevant neuronal activity^83,84^. Higher parietal P300 amplitude is associated with better memory recall and increases with the distinctiveness of an event making an event more likely to be remembered^78,79^. Notably, sleep deprivation reduces P300 amplitude and increases P300 latency while performing an oddball task^80^. In contrast, acute exercise boosts P300 amplitude which corresponds to better cognitive performance^81,82^. Thus, to benefit memory performance, we expected the exercise group to engage P300 mechanism more than the two other groups. To test the abovementioned behavioural and mechanistic hypotheses, we subjected young healthy participants to 30 hours of continuously monitored wakefulness (sleep deprivation (SD)) in the lab. Following this, they were randomized into 20-minute aerobic exercise (EXE), 90-minute nap (NAP) or 20-minute no-intervention control (CON) groups. After the interventions, all the participants were shown images (encoding session) without any instruction of a subsequent memory test. Three days later, participants were tested on their memory for these images (retention session, hereafter referred to as test session) in a Yes-No recognition paradigm by showing them a mix of previously shown images with new ones. Brain activity using 64 channel EEG cap was recorded during both the encoding and test sessions. The brain activity findings discussed hereafter are from the encoding session only.

## Results

The characteristics of the study participants are reported in Table 1.

**Table 1.**
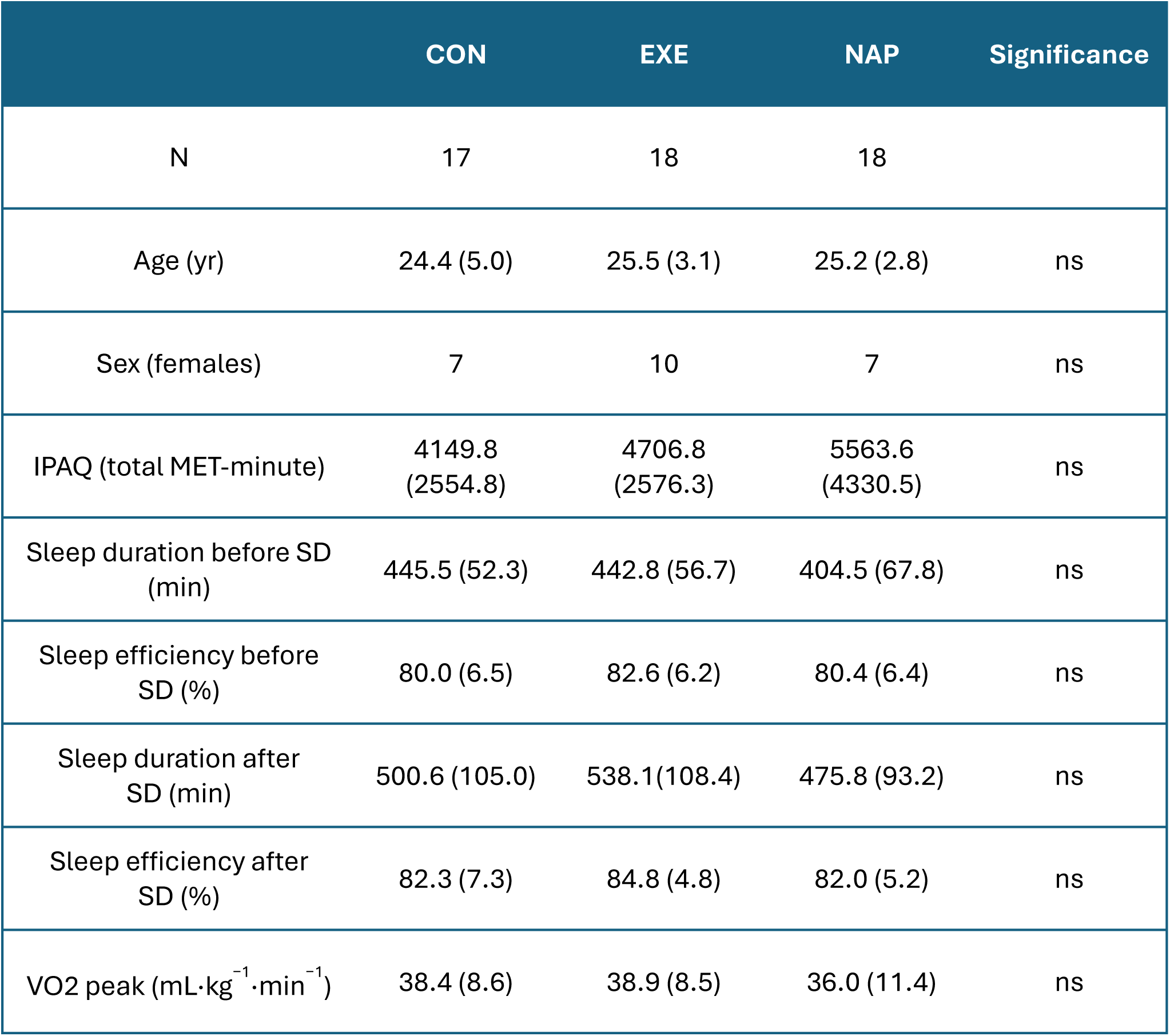
Characteristics of the participants. Data is presented as mean (standard deviation). Reported sleep parameters were analyzed from the actigraphy. ns = non-significant (p>0.05). IPAQ= International Physical Activity Questionnaire; MET = Metabolic equivalent, SD= sleep deprivation. PSQI = Pittsburgh Sleep Quality Index; VO2 peak = Peak volume of oxygen uptake.

### Behavioural findings

Memory for the encoded images after sleep deprivation significantly differed across groups (*F(2,50) = 4.49, p =0.0161*) (Table 2). Post-hoc comparison revealed significantly better memory performance in both the exercise (*p = 0.05, Cohen’s d = 1.0*) and the nap group (*p = 0.0214, Cohen’s d = 0.91*) than the control group, with average improvements of 21% (*MD = 0.098, SE diff = 0.04*) and 23% (*MD= 0.11, SE diff = 0.04*) respectively. Strikingly, memory performance did not significantly differ between exercise and nap group, suggesting similar benefit of these interventions. Both interventions showed large effect sizes (>0.8), underscoring their effectiveness in restoring memory function after sleep loss^85^.

**Table 2.** Behavioural findings. Data is presented as mean (standard deviation). * For non-parametric comparison, median and inter-quartile range is reported. n.a. = post-hoc analyses was not performed. Direction of differences between the groups are reported when group differences are significant. “=” indicates non-significant group differences. Fatigue by SD= fatigue before intervention – fatigue before night of SD (higher value indicates higher fatigue). Fatigue by intervention = fatigue after intervention – fatigue before intervention (negative value indicates decrease in fatigue).

|  | CON | EXE | NAP | p value | Post-hoc group differences |
| --- | --- | --- | --- | --- | --- |
| Memory outcomes |  |  |  |  |  |
| Hit rate | 0.46 (0.11) | 0.56 (0.10) | 0.57 (0.13) | 0.016 | EXE = NAP > CON |
| False alarm rate | 0.17(0.10) | 0.24 (0.12) | 0.19 (0.11) | 0.28 | n.a. |
| Fatigue by SD | 59.41 (31.80) | 52.88(29.52) | 63.72(36.21) | 0.60 | n.a. |
| Fatigue by intervention* | 0.5<br>(-7.0 – 7.25) | -1<br>(-13.0 – 7.75) | -34<br>( -59.0 – -19.25) | <0.0001 | NAP < EXE = CON |

Next, since fatigue affects memory, we compared two types of fatigue across groups: fatigue due to sleep deprivation (fatigue-SD) and fatigue due to interventions (fatigue-int) (Table 2)^86,87^. We found that all groups experienced similar fatigue-SD (*F(2,50) = 0.5, p = 0.60*) and, as expected, differentially experienced fatigue-int (χ²*(2) = 19.76, p<0.0001*). Post-hoc comparisons showed that the nap group experienced significantly lower fatigue-int than exercise (*MD= −12.16, SE diff= 3.41, p=0.0004*) and control (*MD= −13.14, SE diff= 3.34, p < 0.0001*) group, while no significant fatigue-int differences were found between exercise and control group (*MD= −1.34, SE diff= 3.21, p >0.05*). Thus, even after SD, our exercise protocol did not increase fatigue as compared to doing nothing. Notably, despite having similar fatigue levels, the exercise group had better memory performance than the control group.

Further supporting the effect of interventions, we examined whether the interventions increased the hit rate (memory of encoded images) due to enhanced encoding capacity or due to their influence on the response bias. In a yes-no recognition task as reported here, hit rate can be inflated if a participant holds a ‘yes’ bias or a liberal criterion to judge an old image as old^88,89^. However, we found no group differences in the response bias and in fact, the participants held a ‘no’ bias (or a conservative criterion, responding “no” to a previously seen image).

Overall, these findings indicate that both exercising and napping after SD were equally effective in mitigating effects of sleep loss on memory encoding capacity compared to doing nothing, despite a significantly higher fatigue observed in the exercise group than the nap group. To investigate the mechanisms underlying this benefit, we analysed the pre-stimulus tonic brain state and post-stimulus episodic memory EEG markers during the encoding session.

### Electrophysiological findings

#### EEG markers of sleep pressure and fatigue

First, we examined the effects of interventions on neural markers of sleep pressure and fatigue i.e. 0.5-8 Hz SW activity over frontal and task-associated regions respectively during a 1-second pre-stimulus period irrespective of subsequent memory of the images (Fig.2, Table 3). Grand average of the spectral plot within each group is reported in the figure 2. Significant group differences in frontal SW activity were observed (*F(2,39) = 5.69, p = 0.0068*). The nap group, as expected, showed significantly lower frontal SW activity than the control group (*post-hoc comparison, p = 0.0069*), and the exercise (*post-hoc comparison, p = 0.056; post-hoc Student’s t test, p = 0.0224*) indicating the reduced sleep pressure after napping. However, despite exercise being known to increase sleep pressure in normal wakefulness, frontal sleep pressure markers did not differ between exercise and control group after sleep deprivation (*post-hoc comparison, p = 0.58*)^90–93^.

**Fig 1.**
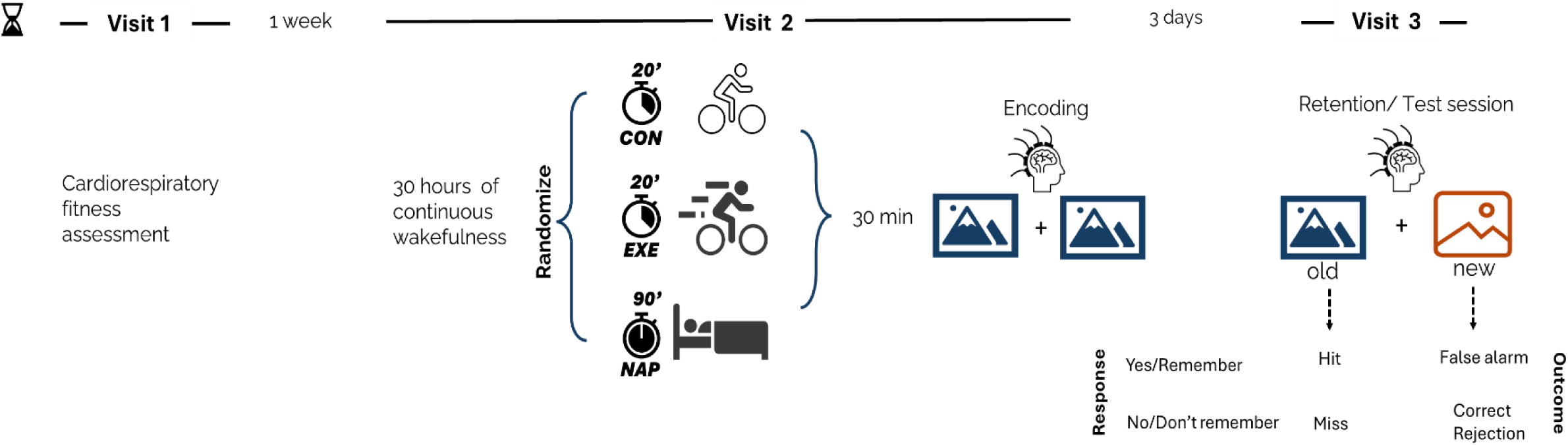
Study design. Each participant did three lab visits spanning over 1.5 weeks. Visit 1: Cardiorespiratory fitness assessment using graded exercise test. Visit 2: 30 hours of lab-monitored sleep deprivation protocol followed by randomization into one of the three groups – CON (20 min sit on bike); EXE (20 min biking at 80% HRmax); NAP (90 min sleep) followed by the encoding session with EEG after 30 min interval; Visit 3: Memory test session (Yes/No paradigm) with EEG.

**Fig 2.**
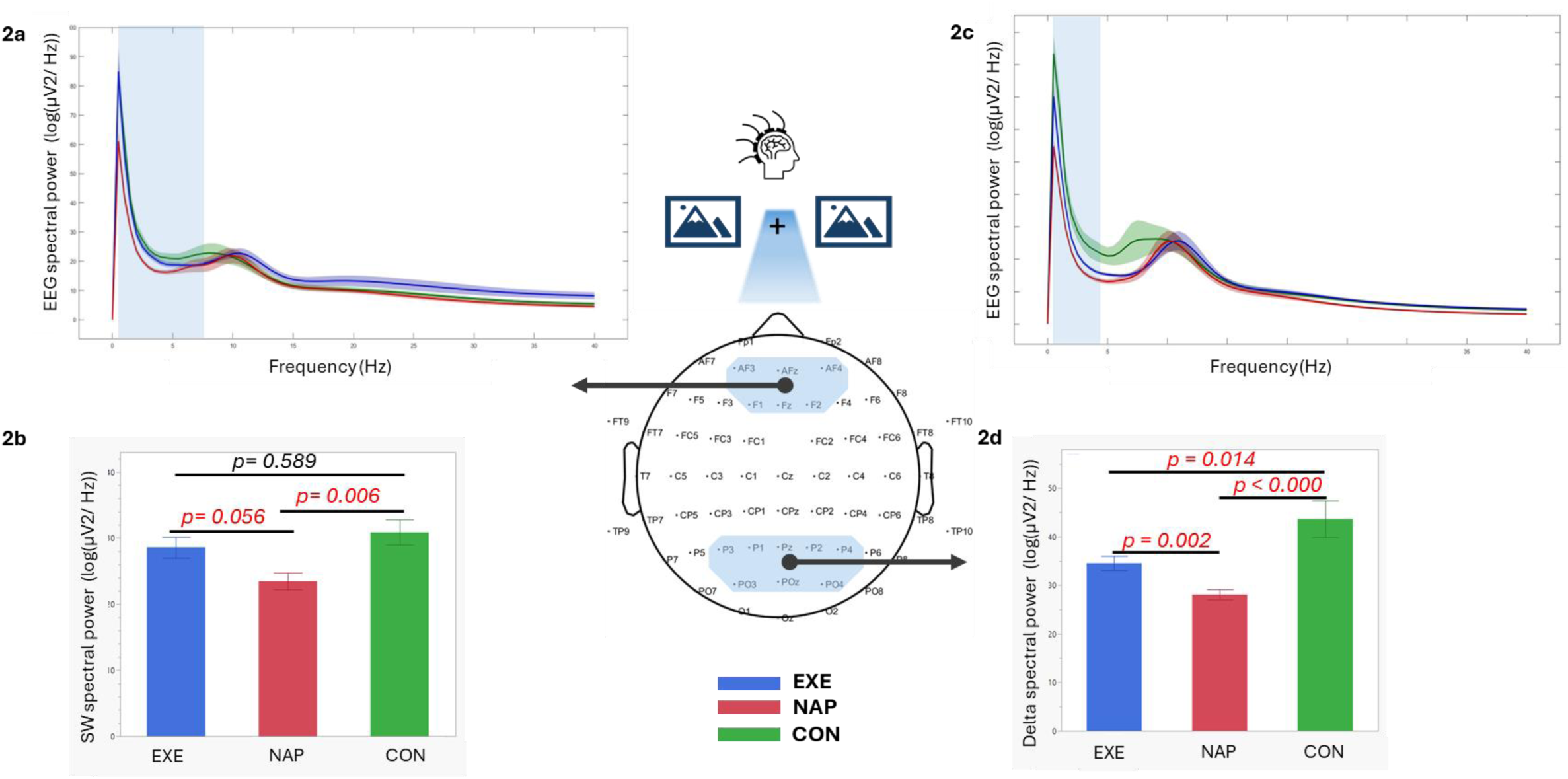
Frontal and task-relevant regional EEG markers of sleep pressure and fatigue are differentially affected by interventions during the pre-stimulus period. Reported are: a) spectral plot and b) group differences in slow wave power (0.5 - 8 Hz) over frontal electrodes c) spectral plot and d) group differences in delta power (0.5 - 4 Hz) over task-relevant regional electrodes. Shaded area around each group’s spectral plot represents standard error of mean. Error bars represent standard error of mean.

**Table 3.** Group differences in the pre-stimulus sleep pressure markers and post-stimulus memory encoding markers in the frontal and task-relevant regions. Mean (SD) values are reported for parametric tests, and Median (IQR) values are reported for non-parametric tests. Direction of group differences are reported when significant. “=” indicates non-significant group differences. SD = standard deviation; IQR = Inter-quartile range; ERSP = Event-related spectral perturbation. * trend of significance for CON – EXE comparison (p = 0.06). ** Mean differences in SW, theta, delta ERSP should be interpreted with caution, as values are normalized to pre-stimulus levels, and pre-stimulus group differences exist (see text). Positive ERSP values signify event-related synchronization; negative ERSP values signify desynchronization. Higher negative values, higher desynchronization, higher positive values, higher synchronization.

|  | Pre-stimulus sleep pressure markers |  |  | Post-Stimulus memory encoding markers |  |  |  |  |
| --- | --- | --- | --- | --- | --- | --- | --- | --- |
|  | Frontal | Task-relevant region |  | Task-relevant region |  |  |  |  |
|  | Slow wave activity | Slow wave activity | Delta activity | Beta ERSP | P300 amplitude | SW ERSP | Theta ERSP | Delta ERSP |
| Groups | Mean (SD) | Median (IQR) | Median (IQR) | Mean (SD) | Mean (SD) | Mean (SD) ** | Mean (SD) ** | Mean (SD) ** |
| CON | 30.85 (6.73) | 26.42 (24.7 9– 32.71) | 38.88 (36.80 – 4.27) | -0.11 (0.07) | 6.72 (2.90) | 0.05 (0.11) | 0.06 (0.16) | 0.04 (0.06) |
| EXE | 28.59 (6.08) | 24.84 (21.34- 26.23) | 35.03 ( 30.25 – 37.13) | -0.16 (0.07) | 5.12 (1.90) | 0.07 (0.06) | 0.09 (0.11) | 0.05 (0.03) |
| NAP | -23.46 (4.96) | 20.19 (17.38 – 23.10) | 28.62 (24.31 – 32.04) | -0.18 (0.04) | 5.54 (1.47) | 0.13 (.011) | 0.16 (0.16) | 0.10 (0.07) |
| p value | 0.0068 | 0.0003 | <0.0001 | 0.0313 | 0.14 | 0.09 | 0.22 | 0.0180 |
| Direction of differences | EXE= CON > NAP | CON>EXE*=NAP | CON> EXE>NAP | NAP > EXE= CON | n/a | n/a | n/a | NAP>EXE=CON |

Additionally, during the same pre-stimulus period, the delta activity (0.5-4 Hz) in the task-relevant region was significantly lower in both the exercise group and nap group compared to the control group *(χ²(2) = 20.82, p<0.0001),* (*post-hoc comparison, MD = −7.6, SE diff = 3.11, p =0.146 and MD = −12.5, SE diff = 3.11, p <0.0001 respectively*) and was lowest in the nap group (*NAP - EXE, MD= −9.733, SE diff= 3.21, p = 0.0025*). These differences persisted for SW activity in the task-relevant region *(χ²(2) = 16.31, p = 0.0003)*, except that the exercise group showed a trend toward lower SW activity than the control group *(post-hoc comparison, p=0.06)* (Fig.2, Table 3). This suggests that the interventions suppressed the neural fatigue in the task-relevant region.

To examine if these group differences reflect local suppression in task-associated regions we compared SW activity in task-relevant and frontal regions within each group. Significant suppression was found only in the exercise (*paired t-test, p = 0.001*) and nap (*Wilcoxon signed-rank paired test, p <0.001*) group, while the control group didn’t show a suppression effect (*paired t-test, p>0.05*). Additionally, across all groups, pre-stimulus SW activity in task-relevant region was significantly correlated with fatigue-int (*Spearman* ρ *= 0.51, p = 0.0005*) and showed a trend of negative correlation with hit rate (*Spearman* ρ *= −0.30, p = 0.053).* These findings suggest that under sleep deprived conditions, these interventions differentially affect brain state: both by altering sleep pressure and by locally suppressing neural fatigue in task-relevant regions contributing to successful memory encoding.

#### EEG markers of episodic memory encoding

Next, we examined if interventions differentially modulated established markers of episodic memory: beta ERD, P300 amplitude and SW ERS in task-relevant regions for all encoding trials, regardless of their subsequent memory (Table 3). Grand averages of the EEG markers within groups are reported in the figure 3. Beta ERD showed significant group differences (*F(2,40) = 3.75, p = 0.0313*). Contrary to our hypothesis, only the nap group showed significantly higher beta ERD than the control group (*post-hoc comparison, p = 0.0270*) with no significant differences between exercise and control or nap groups (*post-hoc comparison, p> 0.05*). Furthermore, despite no difference in comparison to other groups, only in the exercise group, beta ERD significantly correlated with hit rate (*Pearson r = 0.54, p = 0.0356*) and d’ (*Pearson r = 0.60, p = 0.0170*), with higher beta ERD associated with higher hit rate (Fig. 4a).

**Fig. 3.**
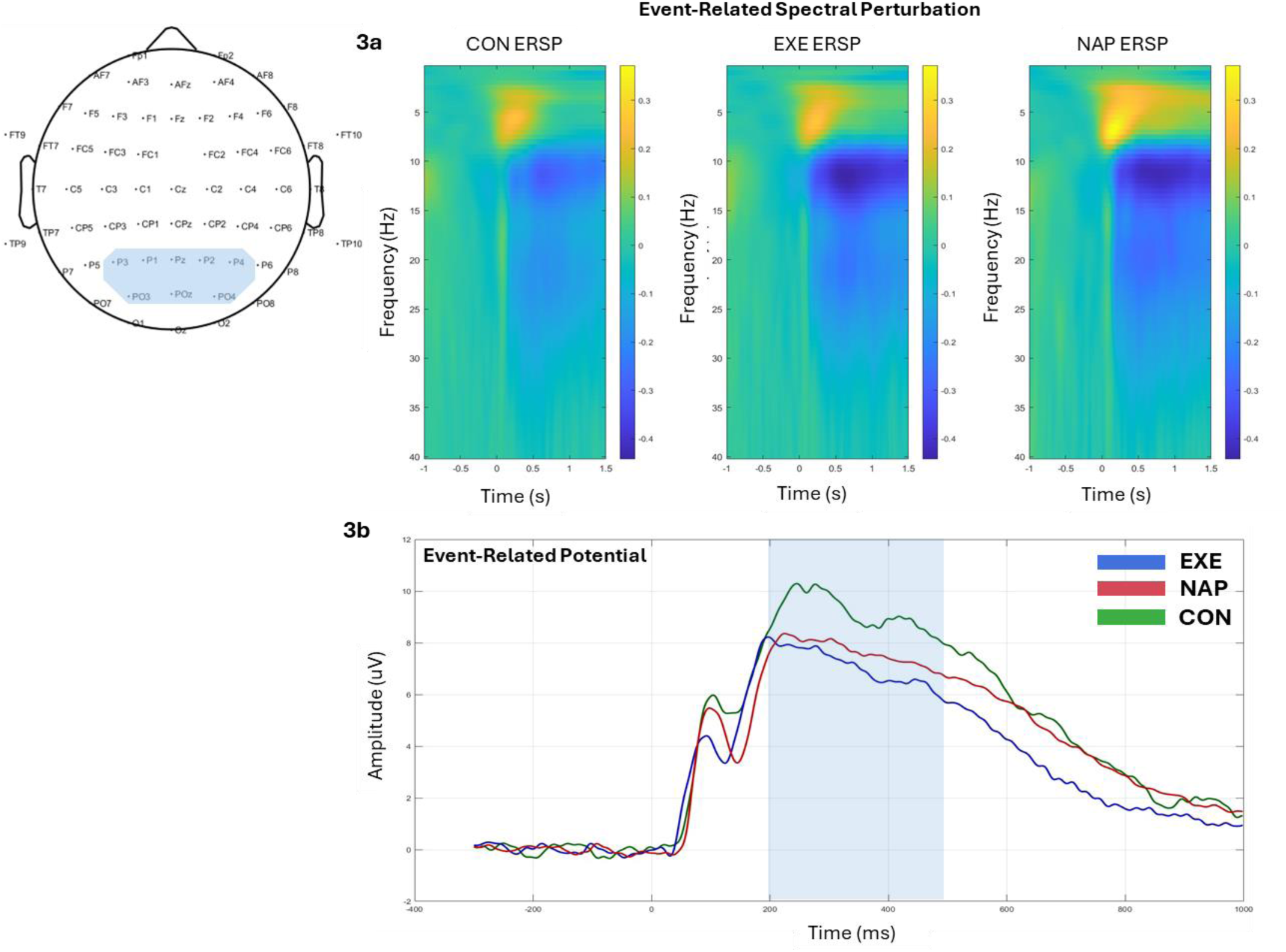
Grand average of EEG markers of episodic memory encoding over task-relevant regional electrodes, shown separately for each group. (a) Grand average event-related spectral perturbation (ERSP) plots. Across all groups, slow-wave synchronization and alpha/beta desynchronization are observed. The x-axis represents time (ms), and the y-axis represents frequency (Hz). (b) Grand average event-related potentials time-locked to image onset and baseline-corrected. The shaded area indicates the time window used for P300 analyses. The x-axis represents time (ms), and the y-axis represents amplitude (µV).

**Fig. 4.**
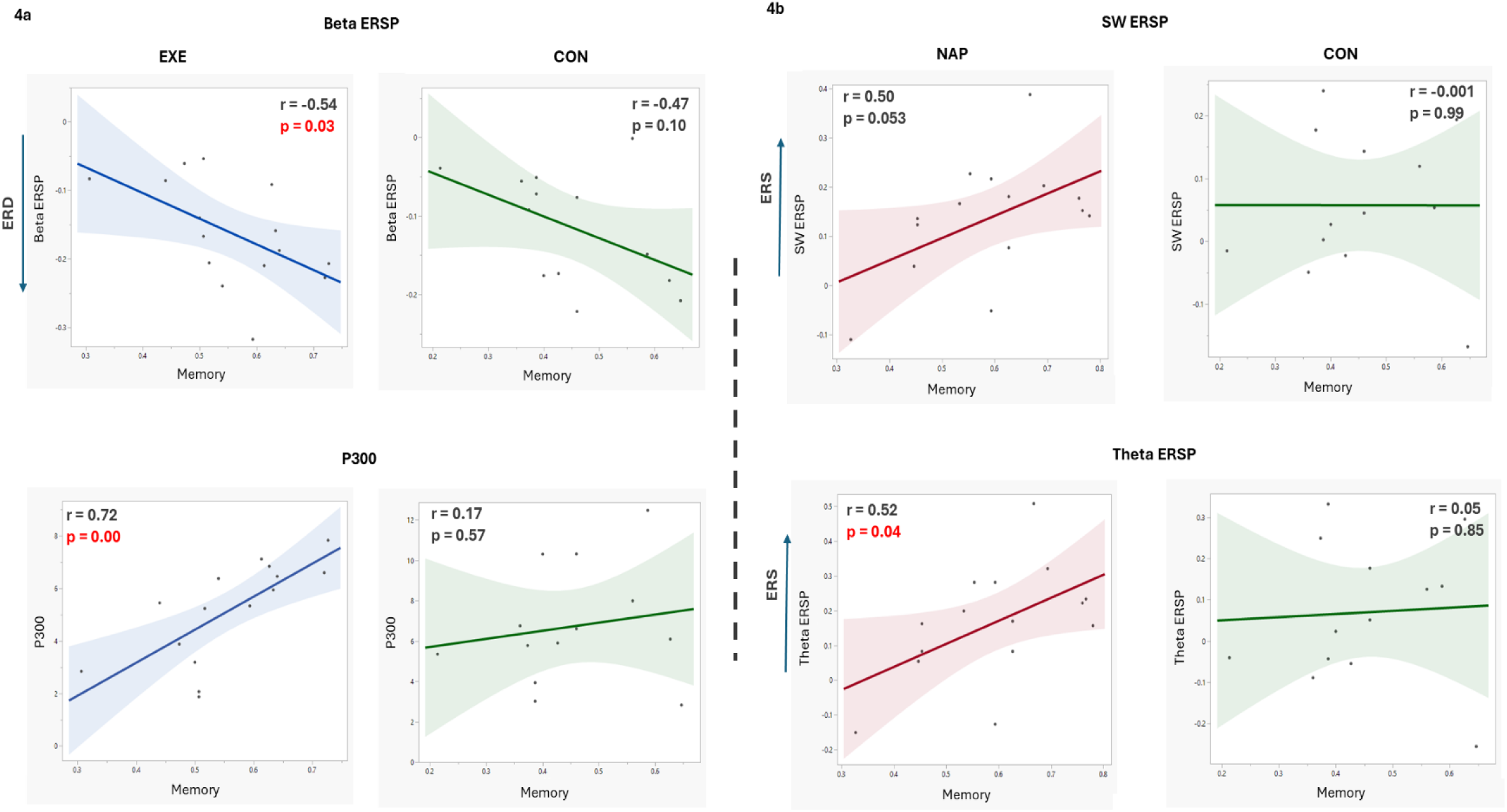
Associations between EEG markers of episodic memory encoding and memory performance within groups. Reported on each plot are the correlation coefficients and corresponding p-values for associations between: 4a) Beta ERSP and P300 amplitude with hit rate in exercise and control groups; 4b) SW ERSP and theta ERSP with hit rate in nap and control groups. Shaded regions represent 95% confidence intervals.

Similarly, surprisingly, no significant group differences for P300 amplitude were found (*F(2,40) = 2.05, p = 0.14*) (although control group showed a trend of higher P300 than the exercise group (*uncorrected student t-test p value = 0.0557*). Again, only in the exercise group did P300 amplitude correlate positively with hit rate (*Pearson r = 0.70, p = 0.0021*). This finding suggests that exercising facilitated the engagement of these mechanisms benefitting memory (Fig. 4a).

Lastly, no significant group differences emerged for theta ERS or slow wave ERS (Table 3). Notably, delta ERS showed significant group differences (*F(2,40)= 4.45, p = 0.0180*) such that the nap group had significantly higher delta ERS than the control group (*post hoc comparison, p=0.023*) and trend of higher delta ERS than the exercise group (*post hoc comparison, p value = 0.0621*). However, these results should be interpreted cautiously as ERS is normalized to pre-stimulus baseline period and can lead to artificial differences in post-stimulus activity. Since we observe the state-dependent pre-stimulus group differences mentioned above (Fig. 2, Table 3), within-group analyses of stimulus-related SW, delta and theta modulation and its association with memory outcomes were considered a better choice for ERS analysis.

Accordingly, all the groups showed stimulus related increase in delta activity (delta ERS) (paired t-test, p <0.05). However, while exercise and nap groups showed a significant increase in theta and SW activity, the control group only showed a trend of this increase (paired t-test, p ∼ 0.05) (extended data Fig.1). Interestingly, only the nap group showed a positive correlation of SW ERS and theta ERS with hit rate (*Pearson r = 0.50, p = 0.05, Pearson r = 0.52, p = 0.043 respectively*) and d’ (*Pearson r = 0.51, p = 0.0470*) (Fig. 4b). This suggests that although stimulus-dependent theta and delta activity increase occurs in all the groups, only the nap group benefitted from this synchronization.

Importantly, supporting the specificity of these interventions to prime the mechanisms of successful memory encoding, the group differences and correlation results with d’ and hit rate also persisted for memory encoding EEG markers during correct trials (extended data, Table 1).

Given that differential neural markers - reported above and in the extended data Table 1-contributed to memory performance, we conducted stepwise multiple regression analyses to identify the strongest predictors of memory outcomes within each group (Table 4). The predictors were selected based on theory and multivariate correlation analyses. Accordingly, within the exercise group, P300 amplitude and beta ERD on correct trials in the task-relevant region emerged as the best neural predictors of hit rate (*F(2,12) = 11.00, p = 0.0019, R² = 0.64, Adj. R² = 0.58, VIF = 1.36, BIC = −29.34*), with greater contribution from P300 (standardized β = 0.66) than from beta ERD (standardized β = 0.22) (Table 4, Fig. 5). Within the nap group, pre-stimulus theta activity and SW ERS on correct trials in the task-relevant region emerged as the best neural predictors of hit rate *F(2,12) = 5.10, p =< 0.0248, R² = 0.46, Adj. R² = 0.37, VIF = 1.5, BIC = −217.41*) with the former contributing more (standardized β = −0.49; than the latter (standardized β = 0.25). However, after removing an influential observation (Cook’s d = 2.36) from the regression analyses, the model fit improved (*F(2,11) = 23.90, p =< 0.0001, R² = 0.81, Adj. R² = 0.77, VIF = 1.25, BIC = −29.68*) with the former contributing more (standardized β = −0.72) than the latter (standardized β = 0.30) (Table 4, Fig. 5). In the control group, none of these predictors significantly predicted the memory outcome.

**Table 4.** Results from the regression analyses predicting hit rate. . All the predictors are computed from the EEG activity over task-relevant region and during correct trials. Reported model summary for the nap excludes an influential observation (Cook’s d = 2.36). Without excluding the model summary is F(2,12) = 5.10, p = 0.0248, Rsq.= 0.46, Rsq. Adj. = 0.37, BIC = −217.41, RMSE = 0.10, VIF = 1.5). SE = Standard error; BIC = Bayesian Information Criterion; RMSE = Root Mean-Squared Error; VIF = Variance Inflation Factor.

| | $\beta$ | SE | Standardized $\beta$ | P value |
| --- | --- | --- | --- | --- |
| **EXE**<br>**F(2,12) = 11.00, p = 0.0019, Rsq. = 0.64, Rsq. Adj. = 0.58, BIC= -29.33, RMSE = 0.07, VIF= 1.36** |  |  |  |  |
| Intercept | 0.28 | 0.06 | 0 | 0.0006 |
| P300 | 0.03 | 0.00 | 0.66 | 0.0062 |
| Beta ERD | -0.35 | 0.31 | -0.22 | 0.2758 |
| **NAP**<br>**F(2,11) = 23.89, p <0.0001, Rsq. = 0.81, Rsq. Adj. = 0.77, BIC= -29.68, RMSE = 0.06, VIF= 1.25** |  |  |  |  |
| Intercept | 1.09 | 0.13 | 0 | 0.0001 |
| Pre-stimulus Theta | -0.04 | 0.00 | -0.72 | 0.0004 |
| SW ERS | 0.58 | 0.28 | 0.30 | 0.0640 |

**Fig. 5.**
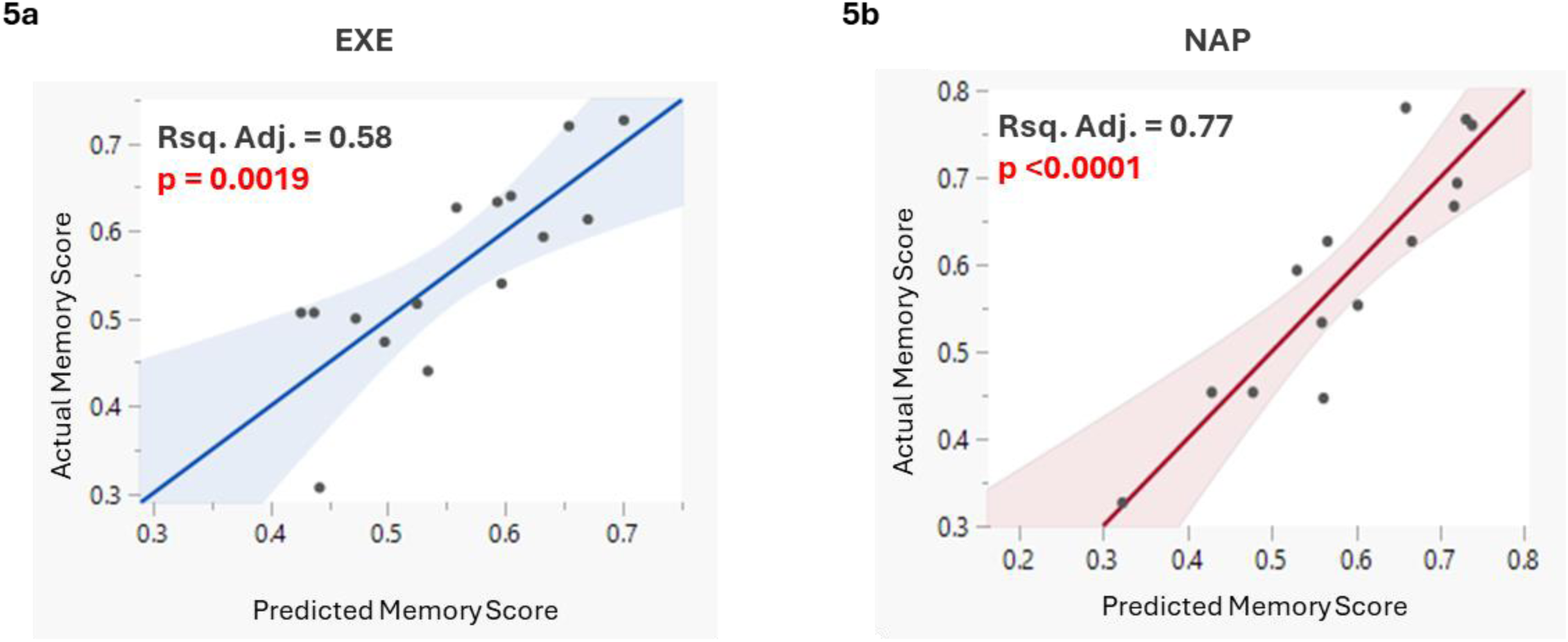
Plot of observed vs. model-predicted memory scores. Reported plots show the relationship between actual memory scores and scores predicted by the regression model in a) EXE group b) NAP group. Memory scores are hit rates. In EXE group, the model includes P300 amplitude and beta-ERD on remembered trials and in the NAP group, the model includes pre-stimulus theta spectral power and SW ERS on remembered trials. Reported plot in the NAP group excludes an influential observation (Cook’s d = 2.36). The regression lines are presented with corresponding 95% confidence intervals.

These findings raise important follow-up questions. Despite comparable levels of sleep pressure, fatigue, and beta ERD in the exercise and control groups, why does only the exercise group benefit from these neural mechanisms? Moreover, although P300 amplitude is positively associated with memory recall in the literature and as reported here, why does the control group, with higher P300 levels than the exercise group, not show improved memory performance? Lastly, given that exercise typically increases P300 amplitude during normal wakefulness, why is this not observed under sleep-deprived conditions? We hypothesized that the elevated P300 levels and lack of correlation of beta ERD with memory in the control group could be explained by other factors.

To answer these exploratory questions, we ran a moderation analysis within the control group and found that P300 levels during remembered trials significantly moderated the relationship between beta ERD on remembered trials and hit rate (*p=0.027*). A Johnson-Neyman analysis revealed that beta ERD only contributed significantly to the memory at lower levels of P300 (extended data Fig.2a). A further investigation revealed that only in the control group, the higher P300 amplitude was significantly associated with greater fatigue-int (*Spearman’s ρ* = 0.58, *p* = 0.036) (extended data Fig.2b). This suggests that, under the sleep-deprived condition, an elevated P300 may reflect compensatory effort due to fatigue rather than benefiting memory. In contrast, the exercise group efficiently engaged beta ERD and P300 (related to memory) mechanisms, not requiring an increase of P300 engagement despite similar levels of fatigue.

## Discussion

This study found that both exercising and napping are equally effective in mitigating the detrimental impact of 30 hours of continuous wakefulness on episodic memory performance in healthy individuals. Moreover, we found that the brain mechanisms significantly contributing to memory performance were specific to each group. Specifically, in the nap group, the memory performance was supported more so by both the pre-stimulus brain state and fatigue-related neural markers and post-stimulus memory encoding markers and in the exercise group, by post-stimulus memory encoding-related markers. The mechanistic findings add to the literature on the degenerate nature of brain, namely its ability to achieve same function through different pathways which allows brain to adapt to insults such as sleep deprivation^94^.

The behavioural findings of this study have important implications in safety-sensitive occupations where sleep loss is prevalent, and working professionals require optimal episodic memory functioning. While napping in such occupations is recommended to counter performance impairments, many implementation barriers limit its application^27,28,95^. On the other hand, there is a growing momentum to integrate exercise and physical activity interventions in various workplace settings to improve mood, fatigue overall health outcomes. Our study supports the utility of such interventions to benefit episodic memory encoding in safety-sensitive occupations^96–98^. Our findings show that exercise can serve as a non-pharmacological alternative to napping in sleep-deprived conditions and inspire further research to study its effectiveness in real-world settings. Moreover, cardiovascular exercise does not require specialized equipment, making it feasible to test its effects in real-world settings. Importantly, as fatigue is a major concern amongst the safety-sensitive occupations, it is worth highlighting that the exercise group did not show higher fatigue (fatigue-int) than the control group despite both experiencing similar levels of fatigue due to sleep deprivation (fatigue-SD)^26,99,100^. Furthermore, the benefits of both interventions on episodic memory encoding persisted for at least 30 minutes, the interval between the interventions and the encoding session, chosen to minimize sleep inertia in the nap group^101,102^. This further highlights the strength of both interventions.

Previous studies investigating the effects of acute exercise on cognitive decline following sleep loss showed mixed results, although no study investigated its effects on episodic memory. For example, Slutsky et al, found neither detrimental nor beneficial effects of exercise on vigilance and working memory performance after 24 hours of continuous wakefulness^103^. This could be due to two factors: firstly, their cohort did not exhibit a significant impairment in working memory post-SD, suggesting that the sleep loss protocol may not have been taxing enough to observe the differences; secondly, they used low-intensity exercise for 15 min which may not be intense enough to show benefits under sleep-deprived conditions. Other studies showing null or detrimental effects of exercise on cognition involved repeated bouts of exercise throughout the SD protocol which may have caused physiological stress rather than protecting cognitive functions. For example, LeDuc et al observed no benefit on various cognitive functions after 10-minute bouts of treadmill exercise done every 2 hours in army aviators^104^. Similarly, another study involving 20-minute exercise bouts every 2 hours in male students resulted in impaired reaction times and worsened mood compared to a no-exercise control^105^. Studies with extreme endurance events, such as ultramarathons lasting 27 – 44 hours showed decrements in cognitive performance likely reflecting the cumulative effect of sleep loss and continuous strenuous exercise effects^106^. These findings suggest that the lack of effectiveness of exercise on cognitive functions after sleep loss could depend on sleep loss intensity, and exercise intensity and duration. Our study demonstrates that a single bout of 20 minutes of aerobic exercise at 80% HR max is enough to benefit episodic memory after 30 hours of continuous wakefulness. These findings add to the emerging evidence on the benefit of acute exercise intervention and moderate to high levels of physical activity on attention and executive function after 24 hours of sleep loss^107,108^.

One striking observation from our study is that we found significant group differences in hit rates but not in false alarm rates (Table 2). This is consistent with prior studies comparing SD and sleep control groups where the false alarm rates did not significantly differ, even though a greater memory impairment is expected between those groups than in our study, where all the groups are sleep deprived^10,11^. Several factors could account for this finding. Firstly, false alarms depend on participants’ responses to new images shown only during the test session, which was three days after encoding in our study and prior work^109,110^. We intentionally chose a three-day interval to allow full recovery from sleep loss and to minimize residual effects of the interventions on retrieval processes^111^. Thus, we did not expect our interventions to differentially prime retrieval mechanisms or the retrieval strategies employed by the participants on the test session which could cause differences in false alarm rates^112^. Moreover, actigraphy data confirmed similar sleep duration and sleep efficiency during the 3-day recovery period and on the immediate night after sleep deprivation (Table 1). The fact that factors influencing memory were similar across groups during the recovery period may explain the lack of significant differences in false alarm rates in our study and prior studies using this memory paradigm. Secondly, increasing encoding strength may not always lead to a reduction in false memory^113^. Lastly, there is a possibility of a floor effect in false alarms in our sample, which may have contributed to the lack of group differences. This finding reinforces that our interventions were specific in priming memory encoding mechanisms and improving encoding strength, reflected in greater hit rates in intervention groups than the control group.

Beyond translational implications, our findings provide novel mechanistic insights into how the brain responds to these plasticity-modulating interventions under sleep-deprived conditions while performing episodic memory encoding. Importantly, our findings show that the intervention groups engaged different mechanisms which significantly contributed to achieving the same level of episodic memory performance, supporting the concept of systems-level plasticity degeneracy^94^. This underscores the brain’s remarkable adaptive capacity to maintain performance despite being sleep deprived^94,114–116^. Contrary to our expectation from the literature in normal wakefulness, we did not observe higher levels of beta ERD and P300 in the exercise group than the control group^69,81,82^. Instead, the control group showed similar levels of beta ERD and a trend of higher P300 levels than the exercise group. Despite this, only in the exercise group both P300 and beta ERD significantly emerged as the two best predictors explaining the variance in memory performance. This suggests that exercise enabled efficient allocation and utility of the available encoding-related neural resources under the challenge of sleep loss. This finding aligns with the neural efficiency hypothesis which states that high-performing individuals show lower task-relevant brain activation than low-performing individuals due to efficient use of cognitive resources^117,118^.

Prior research in expert athletes has shown that exercise training increases neural efficiency, needing lower energy consumption and reduced ERP amplitude, alpha ERD and cortical activation compared to novice athletes to achieve optimal performance^117,119^. Neural efficiency is important under sleep-deprived conditions as SD poses an energetic challenge to the brain, impairs functional connectivity within task-associated regions, uncouples resting state networks, changes cerebral blood flow, decreases brain’s capacity for beta ERD and increases task-associated P300 levels^5,120,121^. Indeed, the impact of sleep loss, sleep disturbances and disorders on cognition is moderated by an individual’s cognitive reserve, neural efficiency being one of the neural bases of this cognitive reserve^122^. Thus, if control and exercise groups have comparable cognitive reserve due to randomization, we surmise that exercise could mobilize the available resources and allocate those resources for the encoding task without needing to further increase ERD or P300 levels. In contrast, in the control group, beta ERD and an increase in P300 levels could indicate an increased effort or increased mobilization of attentional resources to meet task demands, a finding also observed in aging and neurological conditions where reduced neural efficiency may require greater cortical activation to meet task demands^118,123^. Importantly, the elevated P300 level in the control group did not contribute to memory performance and in contrast, was associated with fatigue (fatigue-int). This highlights that elevation of P300 levels under sleep-deprived conditions may reflect a compensatory mechanism, which may not always be beneficial for task performance.

Next, a conceptual inquiry is warranted for our observation that, although a stimulus-dependent increase in slow wave (0.5-8 Hz) occurred across all groups, slow wave ERS significantly predicted the memory performance only in the nap group. We surmise that this finding can be partly explained by ‘the sleep–wake dependent window of optimal associative plasticity’ or ‘happy-medium’ model proposed by Kuhn et. al.^124^. This model suggests that there exists an optimal window of LTP inducibility across the sleep-wake cycle, below or above which the LTP inducibility is attenuated due to, respectively, insufficient upscaling of synapses (immediately after sleep) or excessive upscaling and saturation of synapses (during extended wakefulness)^124^. We argue that, in line with the synaptic homeostasis hypothesis, in our study, napping facilitated synaptic downscaling to a certain degree and placed the brain in a state within the optimal window of LTP inducibility, renewing memory encoding capacity^9,47^. Thus, in the nap group, the pre-stimulus synaptic strength was lower as compared to the exercise and the control group and a stimulus-dependent SW ERS benefited memory only in this group. In contrast, with already potentiated synapses (i.e. higher pre-stimulus slow wave power) in the exercise and the control group, a further synchronisation of SW placed the brain in a state far from the optimal window of LTP inducibility and thus did not capable of benefitting memory encoding. This could explain why both pre-stimulus brain state and post-stimulus slow wave ERS significantly predicted memory encoding success (i.e., hit rate) only in the nap group.

Next, it is worth discussing the findings showing that pre-stimulus theta activity in the nap group predicted poor memory performance. This contrasts prior studies conducted in normal wakefulness where elevated pre-stimulus theta activity predicted successful encoding in various memory paradigms such as cross-modal associative memory, free word recall task, old-new recognition memory, and retrieval of episodic memories^125–128^. These studies argue that in normal wakefulness, pre-stimulus theta activity reflect attentional readiness, cognitive control, and preparedness to learn incoming information. While under sleep-deprived conditions, the increase in slow oscillation - which includes theta - reflects an increase in sleep pressure that is associated with cognitive lapses^60,129,130^. Supporting this, prior studies in sleep-deprived humans and rats have demonstrated that local slow/theta oscillations were associated with neuronal silencing that preceded cognitive lapses^60,130^. It has been proposed that both: cognition theta and sleep deprivation theta can coexist during tasks performed under sleep-deprived conditions such that an increase in theta could reflect either passive cortical disengagement in which the underlying network or brain area ceases to receive inputs or an active cortical inhibition of task-irrelevant networks^129^. In our study, pre-stimulus theta in task-relevant areas may reflect a cumulative effect of use-dependent fatigue and sleep pressure which might necessitate cortical disengagement (and local sleep) at the expense of episodic learning, explaining the contrasting findings from the literature from normal wakefulness^53,129,131^. This suggests that the cognitive benefit of pre-stimulus theta activity may be state-dependent.

We acknowledge some limitations of this study which pose important opportunities for future research. First, we did not do source localization of slow wave activity in frontal and task-relevant regions before and after stimulus presentation, which could provide deeper mechanistic insights into whether the neural sources of cognition-related and sleep pressure-related theta/delta activity are differentially affected by the interventions. Second, if the benefit of SW ERS only in the nap group indeed stems due to reduced sleep pressure, as we hypothesized, an analysis of slow-wave sleep architecture and its duration during napping intervention would help validate this mechanism. However, we suspect that this may be difficult to confirm due to limited variability in slow-wave sleep duration during the 90-minute nap opportunity following sleep deprivation, because in our study median nap efficiency was 96% (measured by actigraphy). Third, analyzing brain activity during the test day could clarify whether encoding mechanisms during subsequent hits on encoding session influenced neural activity during hits on the test day. Additionally, it would be worthwhile to investigate if interventions differentially affect recollection (context-rich memory) and familiarity (context-free knowing) both of which contribute to the recognition memory performance and are known to have different neural substrates^132^. Lastly, while our study was designed to identify an alternative, non-pharmacological strategy to napping with implications in safety sensitive occupations, we acknowledge that lab-induced acute SD may not fully represent the sleep loss experienced by the personnel in the safety sensitive occupations, who may show compounded effects of stress, circadian rhythm misalignment, and sleep restriction and disturbances amongst other factors. Nonetheless, our study offers a promising first step for future implementation studies at such workplaces.

## Conclusion

In the current sleep-deprived society identifying effective solutions to protect brain health, cognition and overall health is critical given the broader societal and economic consequences. We show that exercise and naps are equally effective in coping with episodic memory impairments after sleep loss, complementing prior evidence on the benefits of acute exercise on cognitive decline after sleep loss. We also demonstrate that different neural mechanisms explained the protective effects of exercise and napping under sleep-deprived state. This study provides a strong impetus to test these interventions in safety-sensitive occupations, where napping opportunities can be limited. Furthermore, an inquiry into integrating exercise into workplace safety policies and fatigue risk management systems where sleep loss is common is warranted. Finally, it is important to recognize that sleep deprivation is a widespread and multi-faceted issue and thus coping with it extends beyond such individual-level solutions to social- and societal-level actions.

## METHODS

### Participants

A total of 54 healthy individuals with the following inclusion criteria were enrolled in the study if they were: 1) between 18 to 35 years of age; 2) without any history of neurological or psychiatric clinical conditions or previous history of sleep disorders; 3) without any contraindication to perform cardiovascular exercise; 4) not taking medications or recreational drugs affecting the nervous system at the time of enrollment; 5) sleeping more than 6 h per night and following a regular sleep routine (i.e., going to bed between 9 pm and 12 am and waking up between 6 am and 10 am); 6) without any psychiatric illnesses. Participants were excluded if they: 1) worked nightshifts; 2) had been on a trans-meridian flight crossing three time zones or more during the month before participation in the study; 3) were consuming more than three cups of stimulants a day (e.g., coffee, tea, energy drinks); 4) used any medication to improve sleep, including melatonin or other over the counter alternatives (e.g., antihistaminic); 5) scored more than 10 on Epworth Sleepiness Scale (ESS). ESS is an 8-item questionnaire assessing the likelihood that one would fall asleep in different situations, indicating excessive daytime sleepiness^133^; iv) scored more than 5 on Pittsburgh Sleep Quality Index (PSQI). PSQI is a 7-item questionnaire pertaining to sleep quality in the past month^134^; v) had an aversion to aerobic exercise assessed using Physical Activity Enjoyment Scale (PACES) (score less than or equal to 16). PACES is a questionnaire to assess participants’ enjoyment of exercise. We created an adapted version of the PACES, containing 8 items that participants are asked to rate on a 7-point scale (e.g., 1 = exercise is no fun at all, 7 = exercise is a lot of fun^135^.The study was approved by the local Ethics Research Board and all participants provided written consent before taking part in the study.

### Experimental Design

The entire study consisted of three lab visits spanning over 1.5 weeks (Fig.1). Throughout the study, participants wore Actiwatch 2® monitor (Respironics, Philips) that measured sleep parameters and physical activity levels. Participants also maintained a sleep diary with questions regarding their sleep and wake-up time, duration, subjective sleep quality, number of awakenings and number of caffeinated drinks. These measures allowed us to compare across groups the baseline sleep duration and efficiency, as well as recovery sleep following sleep deprivation, all of which may influence memory performance.

On the first visit, cardiorespiratory fitness (VO₂ peak) was assessed using a gold-standard graded exercise test (GXT) on a stationary bike. Participants’ maximum heart rate (HRmax) was determined during this test. Given the evidence that higher VO₂ peak benefits episodic memory under sleep-deprived conditions, this test allowed us to ensure comparable fitness levels across all groups^110^. For the following week, participants were instructed to get at least 6 hours of sleep every night with a regular sleep schedule until the next visit to avoid an accumulation of sleep debt beyond our SD protocol. If this sleep schedule was not followed, participants were given 3 more days to adjust their sleep schedule before their SD visit. 48 hours before the second visit, participants were refrained from engaging in vigorous physical activity.

On the second visit, participants remained awake for a total of 30 hours, calculated from the time they woke up. They arrived at the laboratory at 9:00 p.m. to begin the supervised SD protocol, during which the participants were refrained from consuming any caffeinated beverages, engaging in physical activity beyond light stretching or walking, and were limited to a maximum of two hours of screen time. To stay engaged, participants took part in activities such as painting, reading, or playing board games.

Immediately after completing the SD protocol, i.e., after 30 hours of continuous wakefulness, participants were fitted with a 64-channel EEG cap and then randomly assigned to the EXE, NAP, or CON group. The EXE group performed 20 minutes of moderate-intensity aerobic exercise on a stationary bike at their 80% HRmax with a cadence of >60 rpm. Participants began with a 3-minute warm-up at 60% HRmax. The rate of perceived exertion was assessed every minute using the 6-20 Borg scale to evaluate the workload^136^. The NAP group was given a 90-minute nap opportunity in the laboratory’s sleep room. They were awakened 90 minutes from the lights out. The CON group sat on the stationary bike for 20 minutes without pedaling. To alleviate the sleep inertia after napping, known to affect cognition, there was a 30-minute interval between the nap and memory encoding task^102,137,138^. This interval was kept consistent across all the groups, even though the duration of interventions was different.

Fatigue and energy were assessed using visual analog scale at participants’ arrival: 9 pm, every 2 hours during the SD protocol, after the SD protocol and immediately before and after the interventions^139,140^. Fatigue measured at 9:00 p.m. and after 30 hours was used to compute fatigue-SD (i.e. fatigue due to sleep deprivation) and fatigue measures from before and after the interventions were used to compute fatigue-int (i.e., fatigue due to intervention).

### Memory Encoding and Test Sessions

At the conclusion of the 30-hour SD period, participants performed the memory encoding task along with EEG monitoring that lasted ∼13 minutes which was administered in the afternoon. Three days later, on Visit 3, participants underwent a surprise memory test along with EEG monitoring. This episodic memory paradigm is validated to be sensitive to 30 hours of SD^109,110^. The encoding and test tasks were designed using the software Superlab 6 (Cedrus, San Pedro, USA).

For the encoding session, participants viewed a total of 150 neutral and colored images divided into 6 randomized blocks of 25 images each. These images were of landscapes, non-renowned people, scenes, and objects taken from the International Affective Picture System (https://csea.phhp.ufl.edu/Media.html). Each image was displayed for 3 seconds, followed by a 2-second fixation cross. Participants were instructed to make a keypad response (RB-740; Cedrus, San Pedro, USA) between 1 to 4 to indicate the intensity of emotions evoked by that picture (1 = low intensity; 4 = high intensity) on the image. The keypresses confirmed the stimulus viewing and that participants remained awake during the session. Trials without responses were excluded from further analysis.

During the memory test session, participants were shown a mix of 150 “old” images and 75 “new” images (neutral, colored sourced from IAPS). Each image was displayed for 3 seconds, during which participants were required to make a forced-choice response between YES and NO on the keypad response (RB-740; Cedrus, San Pedro, USA). “YES” indicated that the participants recognized the image as “old” (an image presented during Visit 2) and “NO” indicated that participants considered that picture as “new” (an image not presented during Visit 2). These responses resulted in four outcome types: (i) hit/remembered trials: old image correctly judged as old; (ii) miss/forgotten trials: old image incorrectly judged to be new; (iii) correct rejection: new image correctly judged as new; and (iv) false alarm: new image incorrectly judged to be old. Hit rate was used as a measure of memory encoding strength (hits/ (hits + misses), d’ is calculated as z(Hit Rate) − z(False Alarm Rate)^89,109,110^. Response bias/criterion was assessed using the following formula: c= −0.5[z (Hit Rate) + z (False Alarm Rate)]^88,89^.

### EEG Procedure and Analyses

EEG activity was recorded during encoding and test sessions using a 64-channel ActiCap cap (Brain Vision, Munich, Germany) with a sampling rate of 500Hz. Electrodes were arranged according to the 10–20 international system. Impedances were lowered to 5 kOhms by applying conductive gel inserted into electrode holders. EEG signals were referenced online to the FCz electrode. A StimTracker® (Cedrus, San Pedro, USA) light sensor placed on the task-computer screen and the response pad were connected to a trigger box to record an event onto the ongoing EEG recording every time an image appeared on the screen or participant made a response, respectively.

EEG signals were pre-processed offline and analyzed using the EEGlab MATLAB toolbox^141^. Briefly, the following steps were followed: 1) Extra EEG segments were removed; 2)EEG signals were bandpass filtered between 0.5 Hz and 50 Hz; 3) bad channels identified using the clean_artifacts (channel correlation parameter: 0.8, flatline channel detection parameter: on) were removed and interpolated^142^; 4) bad segments were removed by visual inspection; 5)EEG signals at each channel was average-referenced; 6)cleaned continuous data was inputted to Infomax Independent component analysis and muscle and eye artifact components were automatically rejected using the ICAlabel method and confirmed with manual inspection^143,144^; 6) the “image” events recorded on the continuous EEG signal during encoding were then converted into “hit” or “miss” depending on the response provided by the participants during the memory test session; 7) the data was epoched (see below) relative to the “image” and “hit” events; 8) the epoched data for each participant was visually inspected for artifacts.

For ERP analyses, continuous data were epoched from −300ms to 1000ms relative to the “image” event regardless its subsequent memory and relative to “hit” trials and baseline corrected. For P300 analyses, the ERP signal was averaged over 200-500 ms post-stimulus (“image”) presentation and “hit” trials over the task-relevant region. The task-relevant region was determined by visualizing topographical maps of evoked brain activity every 100ms relative to the image presentation across participants. Accordingly, all the outcomes measured within the task-relevant region are averaged across P1, P2, Pz, P3, P4, PO3, POz, PO4 electrodes (extended data Fig. 3). Activity over the frontal region was averaged across AF3, AFz, AF4, F1, Fz, F2 electrodes.

For spectral (sleep pressure and fatigue EEG markers), and event-related spectral perturbation (ERSP) analyses i.e ERD and ERS analyses, continuous EEG signals from all electrodes were transformed into the time-frequency domain using a Morlet wavelet (wave number = 7) in the 0.5 Hz to 40 Hz frequency range whit 0.5 Hz steps. This time-frequency resolved (TFR) signal was then epoched between −1000ms and 1500ms relative to the “image” and “hit” trials.

Pre-stimulus period (−1000 ms to 0 ms) was used to compute sleep pressure and fatigue markers in the delta (0.5-4 Hz), theta (4-8Hz) and SW (0.5-8 Hz) frequency bands. For ERSP analyses, TFR epochs were normalized according to the following formulae:

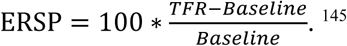

Thus, the computed values post-stimulus indicate an increase in ERSP (ERS) or decrease in ERSP (ERD) relative to the baseline. ERD and ERS values were averaged for “image” and “hit” trials during the post-stimulus period for beta (13-30Hz), and SW (0.5-8 Hz) frequency bands as they are the established memory encoding markers.

### Statistical Analysis

All the extracted behavioral and EEG outcomes were then exported to the JMP® PRO 18.0.2 Software for statistical analyses. For behavioral analyses, data from 18 participants each in the EXE and NAP groups and 17 participants in the CON group were included. One participant from the CON group was excluded due to inattention during the test session. For the EEG analysis, data from 15 participants each in the EXE and NAP groups and 13 participants in the CON group were used. EEG data were excluded for participants who exhibited excessive movement, muscle artifacts, or technical issues.

Normality of distribution within each group for all the variables was determined using Shapiro– Wilk’s Goodness of Fit test. We assessed the group differences in memory and EEG markers using a one-way ANOVA for normally distributed dependent variables or using Wilcoxon / Kruskal-Wallis Tests (Rank Sums) otherwise. When significant, post-hoc comparisons between all group pairs were performed using Tukey-Kramer HSD (normal distribution) or Wilcoxon (non-normal distribution). For within-group analyses, the normality of the difference was determined and accordingly, a paired t-test was used for the normal distribution and Wilcoxon signed-rank test was used for non-normal distribution. Mean and standard deviations are reported for normally distributed variables and median and interquartile range are reported otherwise. Effect sizes were calculated using Cohen’s d formula^85^. Associations were assessed independently for each group. Pearson’s correlation coefficient or Spearman’s rho was used for normally and non-normally distributed variables, respectively. For regression analyses within groups, data points with Cook’s distance values >1 were considered highly influential. Accordingly, one participant from the NAP group was excluded from the regression analyses as the Cook’s d was 2.36. Results before and after excluding this participant are reported. All analyses were performed with two-tailed probability tests with the statistical level (α) set at 0.05.

## Data Availability

All data produced in the present study are available upon reasonable request to the authors

## EXTENDED DATA (FIGURES)

**Fig. 1.**
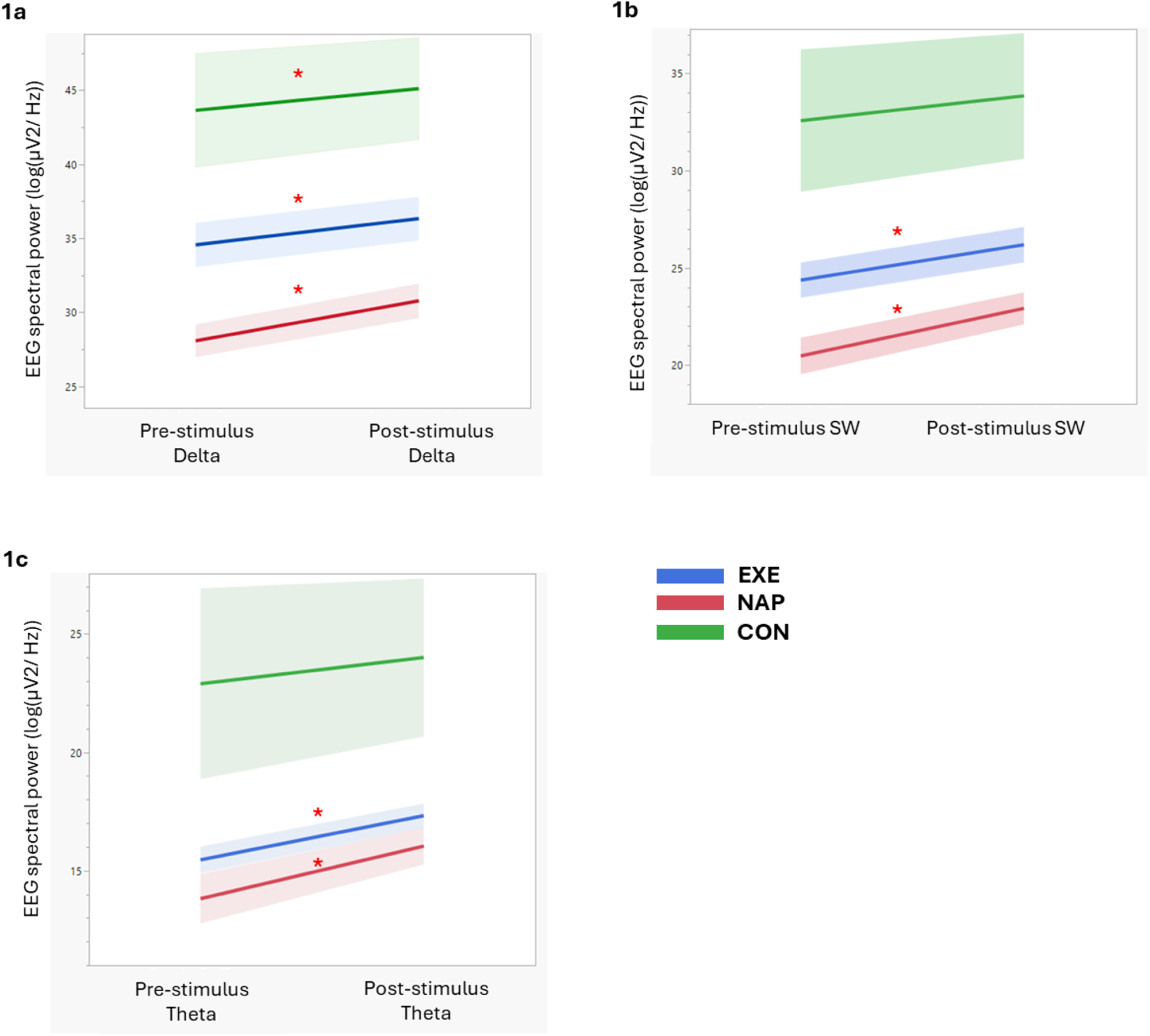
Shown are the three figures resulting from paired t-test analyses performed within three groups between pre-stimulus and post-stimulus a) delta, b) SW and c) theta spectral power. Analyses were done on raw uncorrected spectral power. “*” indicates a significant stimulus dependent increase in spectral power.

**Fig. 2.**
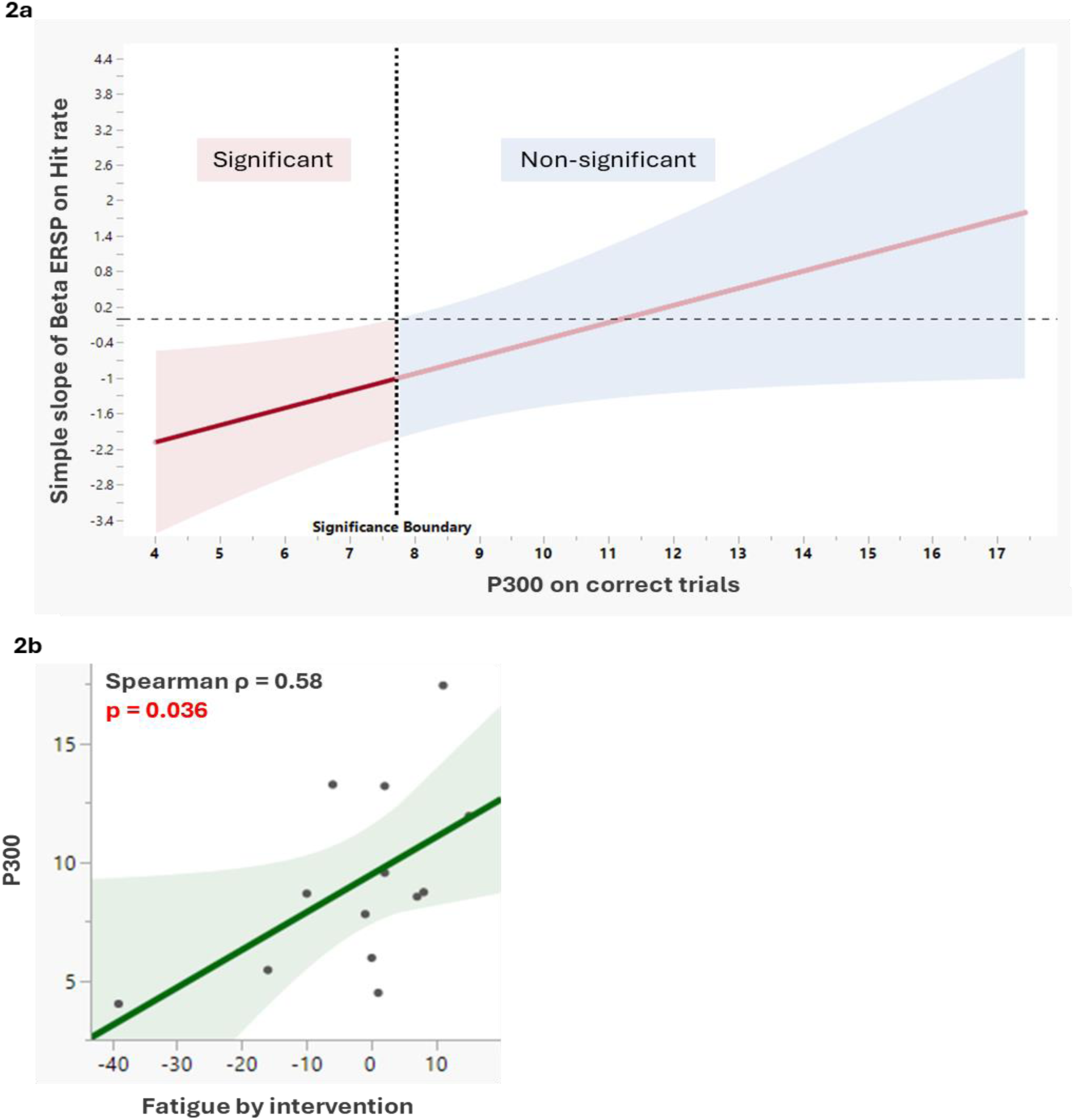
Shown are: a) the Johnson-Neyman plot from the moderation analysis between beta-ERD during remembered trials and hit rate with P300 on remembered trials as a moderator within the CON group. The vertical dashed line is the moderator threshold below which the relationship between beta-ERD during remembered trials and hit rate is significant (p < 0.05). The moderator level at the significant boundary is 7.7. b) the association plot between P300 levels and fatigue-int observed only in the CON group. Negative fatigue values indicate lower fatigue.

**Fig. 3.**
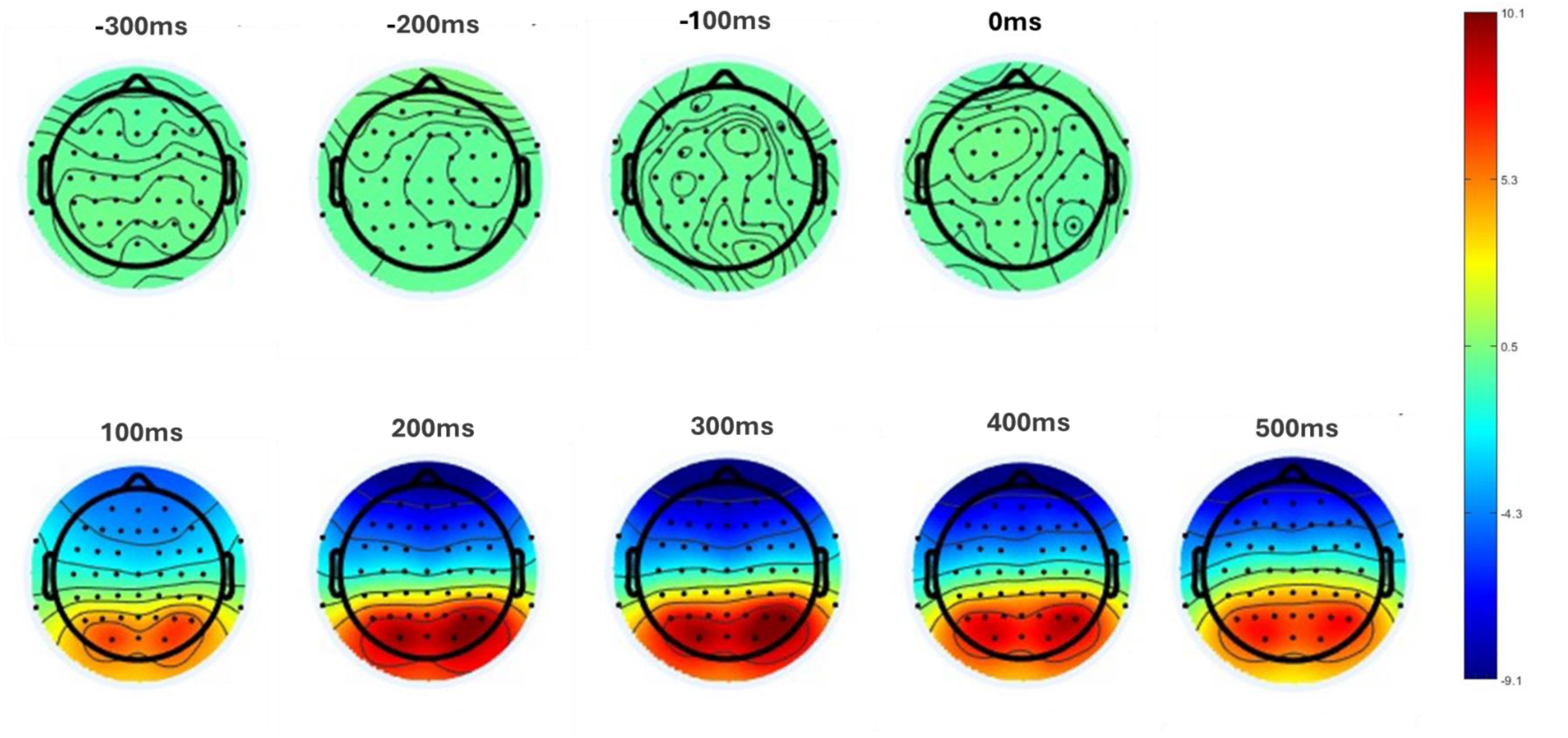
Shown are the topographical time-series plots of evoked potentials from −300 ms to 500 ms relative to stimulus onset, displayed at 100 ms intervals. Parietal-occipital activity emerged following stimulus presentation and thus was identified as the task-relevant region, which is consistent with the literature using a visual stimuli.

## EXTENDED DATA (TABLES)

**Table 1.** Post-stimulus episodic memory encoding markers on correct trials and their correlation with hit rate. Reported are mean (SD) and correlation coefficients of respective encoding markers with hit rate. Direction of group differences are reported when significant. “=” indicates non-significant group differences. SD = standard deviation; ERSP = Event-related spectral perturbation *Mean differences in SW, theta and delta ERSP should be interpreted with caution, as values are normalized to pre-stimulus levels, and pre-stimulus group differences exist (see text). Positive ERSP values signify event-related synchronization; negative ERSP values signify desynchronization. Higher negative values, higher desynchronization, higher positive values, greater synchronization.

| Memory encoding markers | Beta ERSP on correct trials |  | P300 on correct trials |  | SW ERSP on correct trials |  | Theta ERSP on correct trials |  | Delta ERSP on correct trials |  |
| --- | --- | --- | --- | --- | --- | --- | --- | --- | --- | --- |
|  | Correlation coefficient (p value) | Mean (SD) | Correlation coefficient (p value) | Mean (SD) | Correlation coefficient (p value) | Mean (SD)* | Correlation coefficient (p value) | Mean (SD)* | Correlation coefficient (p value) | Mean (SD)* |
| CON | -0.39 (0.17) | -0.09 (0.05) | 0.30 (0.30) | 9.14 (3.92) | 0.32 (0.3) | 0.02 (0.10) | 0.28 (0.34) | 0.02 (0.14) | 0.28 (0.34) | 0.01 (0.05) |
| EXE | -0.57 (0.02) | -0.13 (0.07) | 0.78 (0.000) | 7.02 (2.30) | 0.04 (0.86) | 0.05 (0.04) | 0.05 (0.84) | 0.05 (0.07) | 0.05 (0.84) | 0.05 (0.02) |
| NAP | -0.26 (0.33) | -0.14 (0.03) | -0.05 (0.84) | 7.63 (1.94) | 0.55 (0.03) | 0.10 (0.07) | 0.57 (0.02) | 0.12 (0.1) | 0.57 (0.02) | 0.07 (0.04) |
| p value |  | 0.07 |  | 0.13 |  | 0.02 |  | 0.07 |  | 0.00 |
| Direction of differences |  | n/a |  | n/a |  | NAP> EXE=CON |  | n/a |  | NAP > EXE= CON |

## References

1. Grandner, M.A. (2017). Sleep, Health, and Society. Sleep Med Clin 12, 1–22. 10.1016/j.jsmc.2016.10.012.

2. Akerstedt, T., Fredlund, P., Gillberg, M., and Jansson, B. (2002). Work load and work hours in relation to disturbed sleep and fatigue in a large representative sample. J Psychosom Res 53, 585–588. 10.1016/s0022-3999(02)00447-6.

3. Costa, G. (2015). Sleep deprivation due to shift work. Handb Clin Neurol 131, 437–446. 10.1016/B978-0-444-62627-1.00023-8.

4. Hafner, M., Stepanek, M., Taylor, J., Troxel, W.M., and van Stolk, C. (2017). Why Sleep Matters-The Economic Costs of Insufficient Sleep: A Cross-Country Comparative Analysis. Rand Health Q 6, 11.

5. Krause, A.J., Simon, E.B., Mander, B.A., Greer, S.M., Saletin, J.M., Goldstein-Piekarski, A.N., and Walker, M.P. (2017). The sleep-deprived human brain. Nat Rev Neurosci 18, 404–418. 10.1038/nrn.2017.55.

6. Walker, M.P., and Stickgold, R. (2004). Sleep-dependent learning and memory consolidation. Neuron 44, 121–133. 10.1016/j.neuron.2004.08.031.

7. Berres, S., and Erdfelder, E. (2021). The sleep benefit in episodic memory: An integrative review and a meta-analysis. Psychol Bull 147, 1309–1353. 10.1037/bul0000350.

8. Hokett, E., Arunmozhi, A., Campbell, J., Verhaeghen, P., and Duarte, A. (2021). A systematic review and meta-analysis of individual differences in naturalistic sleep quality and episodic memory performance in young and older adults. Neurosci Biobehav Rev 127, 675–688. 10.1016/j.neubiorev.2021.05.010.

9. Diekelmann, S., and Born, J. (2010). The memory function of sleep. Nat Rev Neurosci 11, 114–126. 10.1038/nrn2762.

10. Yoo, S.S., Hu, P.T., Gujar, N., Jolesz, F.A., and Walker, M.P. (2007). A deficit in the ability to form new human memories without sleep. Nat Neurosci 10, 385–392. 10.1038/nn1851.

11. Ayotte, B., Cristini, J., Lotlikar, M., Parwanta, Z., Cossette, P., Gasparovic, L., Yee-Wong, M., He, Q.Y., Doyon, J., Dal Maso, F., et al. (2023). Does Cardiorespiratory Fitness Protect Memory from Sleep Deprivation? Med Sci Sports Exerc 55, 1632–1640. 10.1249/MSS.0000000000003200.

12. Van Der Werf, Y.D., Altena, E., Schoonheim, M.M., Sanz-Arigita, E.J., Vis, J.C., De Rijke, W., and Van Someren, E.J. (2009). Sleep benefits subsequent hippocampal functioning. Nat Neurosci 12, 122–123. 10.1038/nn.2253.

13. Tulving, E. (2002). Episodic memory: from mind to brain. Annu Rev Psychol 53, 1–25. 10.1146/annurev.psych.53.100901.135114.

14. Biderman, N., Bakkour, A., and Shohamy, D. (2020). What Are Memories For? The Hippocampus Bridges Past Experience with Future Decisions. Trends Cogn Sci 24, 542–556. 10.1016/j.tics.2020.04.004.

15. Addis, D.R., Wong, A.T., and Schacter, D.L. (2007). Remembering the past and imagining the future: common and distinct neural substrates during event construction and elaboration. Neuropsychologia 45, 1363–1377. 10.1016/j.neuropsychologia.2006.10.016.

16. Schacter, D.L., Benoit, R.G., and Szpunar, K.K. (2017). Episodic Future Thinking: Mechanisms and Functions. Curr Opin Behav Sci 17, 41–50. 10.1016/j.cobeha.2017.06.002.

17. Buckner, R.L., Logan, J., Donaldson, D.I., and Wheeler, M.E. (2000). Cognitive neuroscience of episodic memory encoding. Acta Psychol (Amst) 105, 127–139. 10.1016/s0001-6918(00)00057-3.

18. Kohler, S., Moscovitch, M., Winocur, G., and McIntosh, A.R. (2000). Episodic encoding and recognition of pictures and words: role of the human medial temporal lobes. Acta Psychol (Amst) 105, 159–179. 10.1016/s0001-6918(00)00059-7.

19. Folkard, S., and Tucker, P. (2003). Shift work, safety and productivity. Occup Med (Lond) 53, 95–101. 10.1093/occmed/kqg047.

20. Leso, V., Fontana, L., Caturano, A., Vetrani, I., Fedele, M., and Iavicoli, I. (2021). Impact of Shift Work and Long Working Hours on Worker Cognitive Functions: Current Evidence and Future Research Needs. Int J Environ Res Public Health 18. 10.3390/ijerph18126540.

21. Golombek, D.A., Eyre, H., Spiousas, I., Casiraghi, L.P., Hartikainen, K.M., Partonen, T., Pyykko, M., Reynolds, C.F., 3rd, Hynes, W.M., Bassetti, C.L.A., et al. (2025). Sleep Capital: Linking Brain Health to Wellbeing and Economic Productivity Across the Lifespan. Am J Geriatr Psychiatry 33, 92–106. 10.1016/j.jagp.2024.07.011.

22. Hale, L., Troxel, W., and Buysse, D.J. (2020). Sleep Health: An Opportunity for Public Health to Address Health Equity. Annu Rev Public Health 41, 81–99. 10.1146/annurev-publhealth-040119-094412.

23. Hale, L., and Hale, B. (2010). Treat the source not the symptoms: why thinking about sleep informs the social determinants of health. Health Educ Res 25, 395–400. 10.1093/her/cyq027.

24. Newbury, C.R., Crowley, R., Rastle, K., and Tamminen, J. (2021). Sleep deprivation and memory: Meta-analytic reviews of studies on sleep deprivation before and after learning. Psychol Bull 147, 1215–1240. 10.1037/bul0000348.

25. Amin, M.M., Graber, M., Ahmad, K., Manta, D., Hossain, S., Belisova, Z., Cheney, W., Gold, M.S., and Gold, A.R. (2012). The effects of a mid-day nap on the neurocognitive performance of first-year medical residents: a controlled interventional pilot study. Acad Med 87, 1428–1433. 10.1097/ACM.0b013e3182676b37.

26. Allison, P., Tiesman, H.M., Wong, I.S., Bernzweig, D., James, L., James, S.M., Navarro, K.M., and Patterson, P.D. (2022). Working hours, sleep, and fatigue in the public safety sector: A scoping review of the research. Am J Ind Med 65, 878–897. 10.1002/ajim.23407.

27. Patterson, P.D., Weaver, M.D., Guyette, F.X., and Martin-Gill, C. (2020). Should public safety shift workers be allowed to nap while on duty? Am J Ind Med 63, 843–850. 10.1002/ajim.23164.

28. Geiger-Brown, J., Sagherian, K., Zhu, S., Wieroniey, M.A., Blair, L., Warren, J., Hinds, P.S., and Szeles, R. (2016). CE: Original Research: Napping on the Night Shift: A Two-Hospital Implementation Project. Am J Nurs 116, 26–33. 10.1097/01.NAJ.0000482953.88608.80.

29. Takahashi, M. (2003). The role of prescribed napping in sleep medicine. Sleep Med Rev 7, 227–235. 10.1053/smrv.2002.0241.

30. Ficca, G., Axelsson, J., Mollicone, D.J., Muto, V., and Vitiello, M.V. (2010). Naps, cognition and performance. Sleep Med Rev 14, 249–258. 10.1016/j.smrv.2009.09.005.

31. Leong, R.L.F., Lo, J.C., and Chee, M.W.L. (2022). Systematic review and meta-analyses on the effects of afternoon napping on cognition. Sleep Med Rev 65, 101666. 10.1016/j.smrv.2022.101666.

32. Roig, M., Nordbrandt, S., Geertsen, S.S., and Nielsen, J.B. (2013). The effects of cardiovascular exercise on human memory: a review with meta-analysis. Neurosci Biobehav Rev 37, 1645–1666. 10.1016/j.neubiorev.2013.06.012.

33. Loprinzi, P.D., Roig, M., Tomporowski, P.D., Javadi, A.H., and Kelemen, W.L. (2023). Effects of acute exercise on memory: Considerations of exercise intensity, post-exercise recovery period and aerobic endurance. Mem Cognit 51, 1011–1026. 10.3758/s13421-022-01373-4.

34. Loprinzi, P.D., Blough, J., Crawford, L., Ryu, S., Zou, L., and Li, H. (2019). The Temporal Effects of Acute Exercise on Episodic Memory Function: Systematic Review with Meta-Analysis. Brain Sci 9. 10.3390/brainsci9040087.

35. Loprinzi, P.D., Roig, M., Etnier, J.L., Tomporowski, P.D., and Voss, M. (2021). Acute and Chronic Exercise Effects on Human Memory: What We Know and Where to Go from Here. J Clin Med 10. 10.3390/jcm10214812.

36. Loprinzi, P.D., Ponce, P., and Frith, E. (2018). Hypothesized mechanisms through which acute exercise influences episodic memory. Physiol Int 105, 285–297. 10.1556/2060.105.2018.4.28.

37. Roig, M., Cristini, J., Parwanta, Z., Ayotte, B., Rodrigues, L., de Las Heras, B., Nepveu, J.F., Huber, R., Carrier, J., Steib, S., et al. (2022). Exercising the Sleepy-ing Brain: Exercise, Sleep, and Sleep Loss on Memory. Exerc Sport Sci Rev 50, 38–48. 10.1249/JES.0000000000000273.

38. Mograss, M., Crosetta, M., Abi-Jaoude, J., Frolova, E., Robertson, E.M., Pepin, V., and Dang-Vu, T.T. (2020). Exercising before a nap benefits memory better than napping or exercising alone. Sleep 43. 10.1093/sleep/zsaa062.

39. Brooks, A., and Lack, L. (2006). A brief afternoon nap following nocturnal sleep restriction: which nap duration is most recuperative? Sleep 29, 831–840. 10.1093/sleep/29.6.831.

40. Dinges, D.F., Orne, M.T., Whitehouse, W.G., and Orne, E.C. (1987). Temporal placement of a nap for alertness: contributions of circadian phase and prior wakefulness. Sleep 10, 313–329.

41. Lumley, M., Roehrs, T., Zorick, F., Lamphere, J., and Roth, T. (1986). The alerting effects of naps in sleep-deprived subjects. Psychophysiology 23, 403–408. 10.1111/j.1469-8986.1986.tb00653.x.

42. Takahashi, M., and Arito, H. (2000). Maintenance of alertness and performance by a brief nap after lunch under prior sleep deficit. Sleep 23, 813–819.

43. Stepan, M.E., Altmann, E.M., and Fenn, K.M. (2021). Slow-wave sleep during a brief nap is related to reduced cognitive deficits during sleep deprivation. Sleep 44. 10.1093/sleep/zsab152.

44. Mander, B.A., Santhanam, S., Saletin, J.M., and Walker, M.P. (2011). Wake deterioration and sleep restoration of human learning. Curr Biol 21, R183–184. 10.1016/j.cub.2011.01.019.

45. Antonenko, D., Diekelmann, S., Olsen, C., Born, J., and Molle, M. (2013). Napping to renew learning capacity: enhanced encoding after stimulation of sleep slow oscillations. Eur J Neurosci 37, 1142–1151. 10.1111/ejn.12118.

46. Mednick, S., Nakayama, K., and Stickgold, R. (2003). Sleep-dependent learning: a nap is as good as a night. Nat Neurosci 6, 697–698. 10.1038/nn1078.

47. Tononi, G., and Cirelli, C. (2006). Sleep function and synaptic homeostasis. Sleep Med Rev 10, 49–62. 10.1016/j.smrv.2005.05.002.

48. Vyazovskiy, V.V., Cirelli, C., Pfister-Genskow, M., Faraguna, U., and Tononi, G. (2008). Molecular and electrophysiological evidence for net synaptic potentiation in wake and depression in sleep. Nat Neurosci 11, 200–208. 10.1038/nn2035.

49. Tononi, G., and Cirelli, C. (2014). Sleep and the price of plasticity: from synaptic and cellular homeostasis to memory consolidation and integration. Neuron 81, 12–34. 10.1016/j.neuron.2013.12.025.

50. Finelli, L.A., Baumann, H., Borbely, A.A., and Achermann, P. (2000). Dual electroencephalogram markers of human sleep homeostasis: correlation between theta activity in waking and slow-wave activity in sleep. Neuroscience 101, 523–529. 10.1016/s0306-4522(00)00409-7.

51. Aeschbach, D., Matthews, J.R., Postolache, T.T., Jackson, M.A., Giesen, H.A., and Wehr, T.A. (1997). Dynamics of the human EEG during prolonged wakefulness: evidence for frequency-specific circadian and homeostatic influences. Neurosci Lett 239, 121–124. 10.1016/s0304-3940(97)00904-x.

52. Cajochen, C., Knoblauch, V., Krauchi, K., Renz, C., and Wirz-Justice, A. (2001). Dynamics of frontal EEG activity, sleepiness and body temperature under high and low sleep pressure. Neuroreport 12, 2277–2281. 10.1097/00001756-200107200-00046.

53. Van Dongen, H.P., Belenky, G., and Krueger, J.M. (2011). A local, bottom-up perspective on sleep deprivation and neurobehavioral performance. Curr Top Med Chem 11, 2414–2422. 10.2174/156802611797470286.

54. Hung, C.-S., Sarasso, S., Ferrarelli, F., Riedner, B., Ghilardi, M., Cirelli, C., and Tononi, G. (2013). Local experience-dependent changes in the wake EEG after prolonged wakefulness.

55. Ahlstrom, C., Jansson, S., and Anund, A. (2017). Local changes in the wake electroencephalogram precedes lane departures. J Sleep Res 26, 816–819. 10.1111/jsr.12527.

56. Hung, C.S., Sarasso, S., Ferrarelli, F., Riedner, B., Ghilardi, M.F., Cirelli, C., and Tononi, G. (2013). Local experience-dependent changes in the wake EEG after prolonged wakefulness. Sleep 36, 59–72. 10.5665/sleep.2302.

57. Bernardi, G., Siclari, F., Yu, X., Zennig, C., Bellesi, M., Ricciardi, E., Cirelli, C., Ghilardi, M.F., Pietrini, P., and Tononi, G. (2015). Neural and behavioral correlates of extended training during sleep deprivation in humans: evidence for local, task-specific effects. J Neurosci 35, 4487–4500. 10.1523/JNEUROSCI.4567-14.2015.

58. Quercia, A., Zappasodi, F., Committeri, G., and Ferrara, M. (2018). Local Use-Dependent Sleep in Wakefulness Links Performance Errors to Learning. Front Hum Neurosci 12, 122. 10.3389/fnhum.2018.00122.

59. Doran, S.M., Van Dongen, H.P., and Dinges, D.F. (2001). Sustained attention performance during sleep deprivation: evidence of state instability. Arch Ital Biol 139, 253–267.

60. Nir, Y., Andrillon, T., Marmelshtein, A., Suthana, N., Cirelli, C., Tononi, G., and Fried, I. (2017). Selective neuronal lapses precede human cognitive lapses following sleep deprivation. Nat Med 23, 1474–1480. 10.1038/nm.4433.

61. Otten, L.J., Quayle, A.H., Akram, S., Ditewig, T.A., and Rugg, M.D. (2006). Brain activity before an event predicts later recollection. Nat Neurosci 9, 489–491. 10.1038/nn1663.

62. McCormick, D.A., Nestvogel, D.B., and He, B.J. (2020). Neuromodulation of Brain State and Behavior. Annu Rev Neurosci 43, 391–415. 10.1146/annurev-neuro-100219-105424.

63. Hanslmayr, S., Staresina, B.P., and Bowman, H. (2016). Oscillations and Episodic Memory: Addressing the Synchronization/Desynchronization Conundrum. Trends Neurosci 39, 16–25. 10.1016/j.tins.2015.11.004.

64. Hanslmayr, S., Volberg, G., Wimber, M., Raabe, M., Greenlee, M.W., and Bauml, K.H. (2011). The relationship between brain oscillations and BOLD signal during memory formation: a combined EEG-fMRI study. J Neurosci 31, 15674–15680. 10.1523/JNEUROSCI.3140-11.2011.

65. Hanslmayr, S., Spitzer, B., and Bauml, K.H. (2009). Brain oscillations dissociate between semantic and nonsemantic encoding of episodic memories. Cereb Cortex 19, 1631–1640. 10.1093/cercor/bhn197.

66. Griffiths, B.J., Martin-Buro, M.C., Staresina, B.P., Hanslmayr, S., and Staudigl, T. (2021). Alpha/beta power decreases during episodic memory formation predict the magnitude of alpha/beta power decreases during subsequent retrieval. Neuropsychologia 153, 107755. 10.1016/j.neuropsychologia.2021.107755.

67. Hanslmayr, S., Staudigl, T., and Fellner, M.C. (2012). Oscillatory power decreases and long-term memory: the information via desynchronization hypothesis. Front Hum Neurosci 6, 74. 10.3389/fnhum.2012.00074.

68. Guttesen, A.A.V., Gaskell, M.G., Madden, E.V., Appleby, G., Cross, Z.R., and Cairney, S.A. (2023). Sleep loss disrupts the neural signature of successful learning. Cereb Cortex 33, 1610–1625. 10.1093/cercor/bhac159.

69. Chang, Y.K., Chu, C.H., Wang, C.C., Song, T.F., and Wei, G.X. (2015). Effect of acute exercise and cardiovascular fitness on cognitive function: an event-related cortical desynchronization study. Psychophysiology 52, 342–351. 10.1111/psyp.12364.

70. Klimesch, W., Doppelmayr, M., Russegger, H., and Pachinger, T. (1996). Theta band power in the human scalp EEG and the encoding of new information. Neuroreport 7, 1235–1240. 10.1097/00001756-199605170-00002.

71. Jutras, M.J., and Buffalo, E.A. (2010). Synchronous neural activity and memory formation. Curr Opin Neurobiol 20, 150–155. 10.1016/j.conb.2010.02.006.

72. Axmacher, N., Mormann, F., Fernandez, G., Elger, C.E., and Fell, J. (2006). Memory formation by neuronal synchronization. Brain Res Rev 52, 170–182. 10.1016/j.brainresrev.2006.01.007.

73. Osipova, D., Takashima, A., Oostenveld, R., Fernandez, G., Maris, E., and Jensen, O. (2006). Theta and gamma oscillations predict encoding and retrieval of declarative memory. J Neurosci 26, 7523–7531. 10.1523/JNEUROSCI.1948-06.2006.

74. Toth, B., Boha, R., Posfai, M., Gaal, Z.A., Konya, A., Stam, C.J., and Molnar, M. (2012). EEG synchronization characteristics of functional connectivity and complex network properties of memory maintenance in the delta and theta frequency bands. Int J Psychophysiol 83, 399–402. 10.1016/j.ijpsycho.2011.11.017.

75. Guntekin, B., and Basar, E. (2016). Review of evoked and event-related delta responses in the human brain. Int J Psychophysiol 103, 43–52. 10.1016/j.ijpsycho.2015.02.001.

76. Harmony, T. (2013). The functional significance of delta oscillations in cognitive processing. Front Integr Neurosci 7, 83. 10.3389/fnint.2013.00083.

77. Emily S. Kappenman, S.J.L. (2012). ERP Components: The Ups and Downs of Brainwave Recordings. In The Oxford Handbook of Event-Related Potential Components, S.J.L.E.S. Kappenman, ed. pp. 4–30.

78. Mecklinger, A., and Kamp, S.M. (2023). Observing memory encoding while it unfolds: Functional interpretation and current debates regarding ERP subsequent memory effects. Neurosci Biobehav Rev 153, 105347. 10.1016/j.neubiorev.2023.105347.

79. Amin, H.U., Malik, A.S., Kamel, N., Chooi, W.T., and Hussain, M. (2015). P300 correlates with learning & memory abilities and fluid intelligence. J Neuroeng Rehabil 12, 87. 10.1186/s12984-015-0077-6.

80. Lima, N.C., Kirov, R., and de Almondes, K.M. (2022). Impairment of executive functions due to sleep alterations: An integrative review on the use of P300. Front Neurosci 16, 906492. 10.3389/fnins.2022.906492.

81. Kao, S.C., Chen, F. T., Moreau, D., Drollette, E. S., Amireault, S., Chu, C. H., & Chang, Y. K. (2022). cute effects of exercise engagement on neurocognitive function: a systematic review and meta-analysis on P3 amplitude and latency. International Review of Sport and Exercise Psychology 18, 111–153. 10.1080/1750984X.2022.2155488.

82. Chang, Y.-K. (2016). Acute exercise and event-related potential: Current status and future prospects. In Exercise-Cognition Interaction: Neuroscience Perspectives, T. McMorris, ed. (Elsevier Academic Press), pp. 105–130. 10.1016/B978-0-12-800778-5.00005-0.

83. Polich, J. (2007). Updating P300: an integrative theory of P3a and P3b. Clin Neurophysiol 118, 2128–2148. 10.1016/j.clinph.2007.04.019.

84. Polich, J. (2012). Neuropsychology of P300. In The Oxford Handbook of Event-Related Potential Components, S.J.L.E.S. Kappenman, ed. (Oxford University Press), pp. 160–188. 10.1093/oxfordhb/9780195374148.013.0089.

85. Cohen, J. (1992). A power primer. Psychol Bull 112, 155–159. 10.1037//0033-2909.112.1.155.

86. Lohnas, L.J., Davachi, L., and Kahana, M.J. (2020). Neural fatigue influences memory encoding in the human hippocampus. Neuropsychologia 143, 107471. 10.1016/j.neuropsychologia.2020.107471.

87. Batouli, S.A.H., Alemi, R., Khoshkhouy Delshad, H., and Oghabian, M.A. (2020). The influence of mental fatigue on the face and word encoding activations. Clin Neurol Neurosurg 189, 105626. 10.1016/j.clineuro.2019.105626.

88. Caren M. Rotello, N.A.M. (2007). Response Bias in Recognition Memory. In Psychology of Learning and Motivation, (Academic Press), pp. 61–94. 10.1016/S0079-7421(07)48002-1.

89. Stanislaw, H., and Todorov, N. (1999). Calculation of signal detection theory measures. Behav Res Methods Instrum Comput 31, 137–149. 10.3758/bf03207704.

90. JrLeDuc, P.A., Caldwell, J.A., Jr., and Ruyak, P.S. (2000). The effects of exercise as a countermeasure for fatigue in sleep-deprived aviators. Mil Psychol 12, 249–266. 10.1207/S15327876MP1204_02.

91. Leticia Hosang, E.M., Ségolène M. R. Guérin, Costas I. Karageorghis (2022). Effects of exercise on electroencephalography-recorded neural oscillations: a systematic review. International Review of Sport and Exercise Psychology 17, 926–979. 10.1080/1750984X.2022.2103841.

92. Crabbe, J.B., and Dishman, R.K. (2004). Brain electrocortical activity during and after exercise: a quantitative synthesis. Psychophysiology 41, 563–574. 10.1111/j.1469-8986.2004.00176.x.

93. Basso, J.C., and Suzuki, W.A. (2017). The Effects of Acute Exercise on Mood, Cognition, Neurophysiology, and Neurochemical Pathways: A Review. Brain Plast 2, 127–152. 10.3233/BPL-160040.

94. Albantakis, L., Bernard, C., Brenner, N., Marder, E., and Narayanan, R. (2024). The Brain’s Best Kept Secret Is Its Degenerate Structure. J Neurosci 44. 10.1523/JNEUROSCI.1339-24.2024.

95. Edwards, M.P., McMillan, D.E., and Fallis, W.M. (2013). Napping during breaks on night shift: critical care nurse managers’ perceptions. Dynamics 24, 30–35.

96. Flahr, H., Brown, W.J., and Kolbe-Alexander, T.L. (2018). A systematic review of physical activity-based interventions in shift workers. Prev Med Rep 10, 323–331. 10.1016/j.pmedr.2018.04.004.

97. Conn, V.S., Hafdahl, A.R., Cooper, P.S., Brown, L.M., and Lusk, S.L. (2009). Meta-analysis of workplace physical activity interventions. Am J Prev Med 37, 330–339. 10.1016/j.amepre.2009.06.008.

98. Matsugaki, R., Kuhara, S., Saeki, S., Jiang, Y., Michishita, R., Ohta, M., and Yamato, H. (2017). Effectiveness of workplace exercise supervised by a physical therapist among nurses conducting shift work: A randomized controlled trial. J Occup Health 59, 327–335. 10.1539/joh.16-0125-OA.

99. Cho, H., Brzozowski, S., Arsenault Knudsen, E.N., and Steege, L.M. (2021). Changes in Fatigue Levels and Sleep Measures of Hospital Nurses During Two 12-Hour Work Shifts. J Nurs Adm 51, 128–134. 10.1097/NNA.0000000000000983.

100. Akerstedt, T., and Wright, K.P., Jr. (2009). Sleep Loss and Fatigue in Shift Work and Shift Work Disorder. Sleep Med Clin 4, 257–271. 10.1016/j.jsmc.2009.03.001.

101. Tassi, P., Bonnefond, A., Engasser, O., Hoeft, A., Eschenlauer, R., and Muzet, A. (2006). EEG spectral power and cognitive performance during sleep inertia: the effect of normal sleep duration and partial sleep deprivation. Physiol Behav 87, 177–184. 10.1016/j.physbeh.2005.09.017.

102. Tassi, P., and Muzet, A. (2000). Sleep inertia. Sleep Med Rev 4, 341–353. 10.1053/smrv.2000.0098.

103. Slutsky, A.B., Diekfuss, J.A., Janssen, J.A., Berry, N.T., Shih, C.H., Raisbeck, L.D., Wideman, L., and Etnier, J.L. (2017). The effects of low-intensity cycling on cognitive performance following sleep deprivation. Physiol Behav 180, 25–30. 10.1016/j.physbeh.2017.07.033.

104. LeDuc, P.A., Caldwell, J.A., Jr., and Ruyak, P.S. (2000). The effects of exercise as a countermeasure for fatigue in sleep-deprived aviators.

105. Scott, J.P., McNaughton, L.R., and Polman, R.C. (2006). Effects of sleep deprivation and exercise on cognitive, motor performance and mood. Physiol Behav 87, 396–408. 10.1016/j.physbeh.2005.11.009.

106. Hurdiel, R., Peze, T., Daugherty, J., Girard, J., Poussel, M., Poletti, L., Basset, P., and Theunynck, D. (2015). Combined effects of sleep deprivation and strenuous exercise on cognitive performances during The North Face(R) Ultra Trail du Mont Blanc(R) (UTMB(R)). J Sports Sci 33, 670–674. 10.1080/02640414.2014.960883.

107. Kojima, S., Abe, T., Morishita, S., Inagaki, Y., Qin, W., Hotta, K., and Tsubaki, A. (2020). Acute moderate-intensity exercise improves 24-h sleep deprivation-induced cognitive decline and cerebral oxygenation: A near-infrared spectroscopy study. Respir Physiol Neurobiol 274, 103354. 10.1016/j.resp.2019.103354.

108. Tongtong Wei, F.G., Weilong Ma (2025). Physical Activity Attenuates the Adverse Effects of HRV and Cognitive Performance after 24h Sleep Deprivation in Male University Students. ICSTPA ′24: Proceedings of the 2024 International Conference on Sports Technology and Performance Analysis, 365–371. 10.1145/3723936.372399.

109. Yoo, S.-S., Hu, P.T., Gujar, N., Jolesz, F.A., and Walker, M.P. (2007). A deficit in the ability to form new human memories without sleep.

110. Ayotte, B., Cristini, J., Lotlikar, M.S., Parwanta, Z., Cosette, P., Gasparovic, L., Yee-Wong, M., He, Q.Y., Doyon, J., Dal Maso, F., Carrier, J., Steib, S., Robertson, E., Roig, M Does cardio-respiratory fitness protect memory from sleep deprivation?. Journal of sleep research.

111. Chai, Y., Fang, Z., Yang, F.N., Xu, S., Deng, Y., Raine, A., Wang, J., Yu, M., Basner, M., Goel, N., et al. (2020). Two nights of recovery sleep restores hippocampal connectivity but not episodic memory after total sleep deprivation. Sci Rep 10, 8774. 10.1038/s41598-020-65086-x.

112. Chad S Dodson, D.L.S. (2002). When False Recognition Meets Metacognition: The Distinctiveness Heuristic. Journal of Memory and Language 46, 782–803. 10.1006/jmla.2001.2822.

113. Reagh, Z.M., and Yassa, M.A. (2014). Repetition strengthens target recognition but impairs similar lure discrimination: evidence for trace competition. Learn Mem 21, 342–346. 10.1101/lm.034546.114.

114. Drummond, S.P., Brown, G.G., Gillin, J., Stricker, J.L., Wong, E.C., and Buxton, R.B. (2000). Altered brain response to verbal learning following sleep deprivation.

115. Drummond, S.P., Brown, G.G., Salamat, J.S., and Gillin, J.C. (2004). Increasing task difficulty facilitates the cerebral compensatory response to total sleep deprivation. Sleep 27, 445–451.

116. Drummond, S.P., Meloy, M.J., Yanagi, M.A., Orff, H.J., and Brown, G.G. (2005). Compensatory recruitment after sleep deprivation and the relationship with performance. Psychiatry Res 140, 211–223. 10.1016/j.pscychresns.2005.06.007.

117. Li, L., and Smith, D.M. (2021). Neural Efficiency in Athletes: A Systematic Review. Front Behav Neurosci 15, 698555. 10.3389/fnbeh.2021.698555.

118. Barulli, D., and Stern, Y. (2013). Efficiency, capacity, compensation, maintenance, plasticity: emerging concepts in cognitive reserve. Trends Cogn Sci 17, 502–509. 10.1016/j.tics.2013.08.012.

119. Babiloni, C., Marzano, N., Infarinato, F., Iacoboni, M., Rizza, G., Aschieri, P., Cibelli, G., Soricelli, A., Eusebi, F., and Del Percio, C. (2010). “Neural efficiency” of experts’ brain during judgment of actions: a high-resolution EEG study in elite and amateur karate athletes. Behav Brain Res 207, 466–475. 10.1016/j.bbr.2009.10.034.

120. Scharf, M.T., Naidoo, N., Zimmerman, J.E., and Pack, A.I. (2008). The energy hypothesis of sleep revisited. Prog Neurobiol 86, 264–280. 10.1016/j.pneurobio.2008.08.003.

121. Mackiewicz, M., Shockley, K.R., Romer, M.A., Galante, R.J., Zimmerman, J.E., Naidoo, N., Baldwin, D.A., Jensen, S.T., Churchill, G.A., and Pack, A.I. (2007). Macromolecule biosynthesis: a key function of sleep. Physiol Genomics 31, 441–457. 10.1152/physiolgenomics.00275.2006.

122. Balsamo, F., Berretta, E., Meneo, D., Baglioni, C., and Gelfo, F. (2024). The Complex Relationship between Sleep and Cognitive Reserve: A Narrative Review Based on Human Studies. Brain Sci 14. 10.3390/brainsci14070654.

123. Dunst, B., Benedek, M., Jauk, E., Bergner, S., Koschutnig, K., Sommer, M., Ischebeck, A., Spinath, B., Arendasy, M., Buhner, M., et al. (2014). Neural efficiency as a function of task demands. Intelligence 42, 22–30. 10.1016/j.intell.2013.09.005.

124. Kuhn, M., Wolf, E., Maier, J.G., Mainberger, F., Feige, B., Schmid, H., Bürklin, J., Maywald, S., Mall, V., Jung, N.H., et al. (2016). Sleep recalibrates homeostatic and associative synaptic plasticity in the human cortex. Nat Commun 7, 12455. 10.1038/ncomms12455.

125. Guderian, S., Schott, B.H., Richardson-Klavehn, A., and Duzel, E. (2009). Medial temporal theta state before an event predicts episodic encoding success in humans. Proc Natl Acad Sci U S A 106, 5365–5370. 10.1073/pnas.0900289106.

126. Merkow, M.B., Burke, J.F., Stein, J.M., and Kahana, M.J. (2014). Prestimulus theta in the human hippocampus predicts subsequent recognition but not recall. Hippocampus 24, 1562–1569. 10.1002/hipo.22335.

127. Addante, R.J., Watrous, A.J., Yonelinas, A.P., Ekstrom, A.D., and Ranganath, C. (2011). Prestimulus theta activity predicts correct source memory retrieval. Proc Natl Acad Sci U S A 108, 10702–10707. 10.1073/pnas.1014528108.

128. Ostrowski, J., and Rose, M. (2024). Increases in pre-stimulus theta and alpha oscillations precede successful encoding of crossmodal associations. Sci Rep 14, 7895. 10.1038/s41598-024-58227-z.

129. Snipes, S., Krugliakova, E., Meier, E., and Huber, R. (2022). The Theta Paradox: 4-8 Hz EEG Oscillations Reflect Both Sleep Pressure and Cognitive Control. J Neurosci 42, 8569–8586. 10.1523/jneurosci.1063-22.2022.

130. Vyazovskiy, V.V., Olcese, U., Hanlon, E.C., Nir, Y., Cirelli, C., and Tononi, G. (2011). Local sleep in awake rats. Nature 472, 443–447. 10.1038/nature10009.

131. Siclari, F., and Tononi, G. (2017). Local aspects of sleep and wakefulness. Curr Opin Neurobiol 44, 222–227. 10.1016/j.conb.2017.05.008.

132. Kwon, S., Rugg, M.D., Wiegand, R., Curran, T., and Morcom, A.M. (2023). A meta-analysis of event-related potential correlates of recognition memory. Psychon Bull Rev 30, 2083–2105. 10.3758/s13423-023-02309-y.

133. Johns, M.W. (1991). A new method for measuring daytime sleepiness: the Epworth sleepiness scale. Sleep 14, 540–545. 10.1093/sleep/14.6.540.

134. Buysse, D.J., Reynolds, C.F., 3rd, Monk, T.H., Berman, S.R., and Kupfer, D.J. (1989). The Pittsburgh Sleep Quality Index: a new instrument for psychiatric practice and research. Psychiatry Res 28, 193–213. 10.1016/0165-1781(89)90047-4.

135. Kendzierski, D., DeCarlo, K. J. (1991). Physical activity enjoyment scale: Two validation studies. Journal of Sport & Exercise Psychology.

136. Borg, G. (1970). Perceived exertion as an indicator of somatic stress. Scand J Rehabil Med.

137. Trotti, L.M. (2017). Waking up is the hardest thing I do all day: Sleep inertia and sleep drunkenness. Sleep Med Rev 35, 76–84. 10.1016/j.smrv.2016.08.005.

138. Kovac, K., Ferguson, S.A., Paterson, J.L., Aisbett, B., Hilditch, C.J., Reynolds, A.C., and Vincent, G.E. (2020). Exercising Caution Upon Waking-Can Exercise Reduce Sleep Inertia? Front Physiol 11, 254. 10.3389/fphys.2020.00254.

139. Lee, K.A., Hicks, G., and Nino-Murcia, G. (1991). Validity and reliability of a scale to assess fatigue. Psychiatry Res 36, 291–298. 10.1016/0165-1781(91)90027-m.

140. Brunet, J.F., Dagenais, D., Therrien, M., Gartenberg, D., and Forest, G. (2017). Validation of sleep-2-Peak: A smartphone application that can detect fatigue-related changes in reaction times during sleep deprivation. Behav Res Methods 49, 1460–1469. 10.3758/s13428-016-0802-5.

141. Delorme, A., and Makeig, S. (2004). EEGLAB: an open source toolbox for analysis of single-trial EEG dynamics including independent component analysis. J Neurosci Methods 134, 9–21. 10.1016/j.jneumeth.2003.10.009.

142. Kothe, C.A., and Scott Makeig (2013). BCILAB: a platform for brain-computer interface development. Journal of neural engineering 10. 10.1088/1741-2560/10/5/056014.

143. Luca Pion-Tonachini, K.K.-D., Scott Makeig (2019). ICLabel: An automated electroencephalographic independent component classifier, dataset, and website. NeuroImage 198, 181–197. 10.1016/j.neuroimage.2019.05.026.

144. Bell, A.J., and T J Sejnowski (1995). An information-maximization approach to blind separation and blind deconvolution. Neural computation 7, 1129–1159. 10.1162/neco.1995.7.6.1129.

145. G Pfurtscheller, A.A. (1977). Event-related cortical desynchronization detected by power measurements of scalp EEG. Electroencephalography and Clinical Neurophysiology 42, 817–826. 10.1016/0013-4694(77)90235-8.

